# The Impact of a Polyphenol-Rich Supplement on Epigenetic and Cellular Markers of Immune Age: A Pilot Clinical Study

**DOI:** 10.1101/2024.07.05.24309915

**Authors:** Austin Perlmutter, Jeffrey S. Bland, Arti Chandra, Sonia S. Malani, Ryan Smith, Tavis L. Mendez, Varun B. Dwaraka

## Abstract

Age-related alterations in immune function are believed to increase risk for a host of age-related diseases leading to premature death and disability. Programming of the immune system by diet, lifestyle, and environmental factors occurs across the lifespan and influences both makeup and function of the immune system. This programming is believed to act in large part through epigenetic modification.

One group of dietary molecules linked to generally healthier aging and relative immune resilience and homeostasis are polyphenols, which are consumed by humans primarily in the form of plant foods. While polyphenols are widely distributed throughout the plant and fungal kingdoms, certain foods are known to possess distinctive and relatively higher levels of these compounds. One such food is Tartary buckwheat (*fagopyrum tataricum*), an ancient seed historically prized for its health benefits. It is suggested that the specific composition of polyphenols found in foods like Tartary buckwheat may lead to a unique impact on longevity-related physiological pathways that could be interrogated through immune and epigenetic analyses.

The objective of this study was to investigate the epigenetic effects on peripheral immune cells in healthy individuals of a standardized polyphenol concentrate based on naturally occurring nutrients in Tartary buckwheat. A pilot clinical trial was designed to test the effects of consuming 90 days of this concentrate on immune cell epigenetic methylation patterns and immune cell phenotypes in 50 healthy male (40%) and female (60%) participants aged 18-85 years using epigenetic age clocks and deconvolution methods. Analysis revealed significant intervention-related changes in multiple epigenetic age clocks and immune markers as well as population-wide alterations in gene ontology (GO) pathways related to longevity and immunity. This study provides previously unidentified insights into the immune, longevity and epigenetic effects of consumption of polyphenol-rich plants and generates additional support for health interventions built around historically consumed plants like Tartary buckwheat while offering compelling opportunities for additional research.

## Background

In the last century, in high-income countries, national life expectancy has steadily risen. Concurrent with a longer lifespan, there has been a rapid rise in the prevalence of chronic conditions including cardiovascular diseases, diabetes, cancers, chronic respiratory diseases (e.g., asthma and chronic obstructive pulmonary disease) (Kontis et al., 2017). These represent significant contributors to morbidity and mortality as well as unsustainable increases in healthcare costs. Thus, increasing the number of healthy years lived (i.e., health span) remains a challenge despite the increase in lifespan.

Biological age is described as a local or organismal rate of cellular aging and has emerged as a superior predictor risk for disease, mortality, and morbidity when compared to chronological age. Biological aging is most often measured through evaluation of epigenetic markers, most commonly methylation of the cytosine-guanine (CpG) islands in DNA. Biological aging and epigenetic aging are often used interchangeably. Notably, many epigenetic testing approaches utilize DNA from peripheral immune cells, which allows for simultaneous assessment of immune-related epigenetic markers that change with age. Algorithms using epigenetic methylation data such as GrimAge, PhenoAge, and Dunedin Pace have been applied to the determination of biological age of immune cells (Ray and Yung, 2018). It has been found that a higher biological age of the immune system represents a significant risk factor for all-cause mortality and reduction in longevity (Ahuja et al., 2023; Karagiannis et al., 2023; Nie et al., 2022).

Shifts in immune cell subsets, cytokines, and functions are seen in chronic disease states ranging from cardiovascular disease to dementia. In addition, research has progressively focused on the role of the immune system in the aging process itself. Pharmaceutical and non-pharmaceutical approaches have been proposed as potential avenues for ameliorating the putative damage from immune aging (i.e., immunosenescence). The combined measurement of epigenetic aging metrics and immune-specific measurements may, therefore, provide considerable insights into one’s risk for disease and disability.

A state of relative immunosenescence (age-related changes in immune system makeup and function) may contribute to chronic disease via the generation of senescent cells– cells outside the cell replicative cycle due to stress or other insults (Liu et al., 2023). While senescence appears to play a key role in tumor suppression, senescent cells can nonetheless remain metabolically active and produce and secrete a wide range of immunologically active molecules, including inflammatory mediators. The development of senescent cells that produce inflammatory mediators has been termed the senescence-associated secretory phenotype (SASP) and is thought to contribute to the higher levels of inflammation that are often seen with advancing chronological and biological age.

Research on interventions to slow the rate of biological aging, including targeting pathways of immunosenescence has primarily been conducted in cell and animal models. Several recent studies have examined the effect of lifestyle and pharmaceutical interventions on biological aging in humans as well. For example, associations between nutritional intervention, or diet and lifestyle, with epigenetic aging have been reported (Gensous et al., 2020; Fitzgerald et al., 2021). Food-related molecules have been proposed to play an outsized protective role in slowing or potentially reversing epigenetic aging. Among the best-studied health-promoting dietary nutrients are polyphenols, non-caloric plant-derived compounds which have a wide variety of proposed effects in humans. Indeed, consumption of a polyphenol-rich beverage was found to correlate with epigenetic changes in immune cells of dyslipidemic humans in a recent study (Stojković et al., 2021). A recent randomized controlled trial (Hoffmann et al., 2023) additionally demonstrated epigenetic effects linked to consumption of a polyphenol-enriched Mediterranean diet proposed to have effects on the immune response.

Several lines of research have more explicitly examined slowing or reversing immunosenescence. Here again, polyphenols have been proposed to play a potential role, and in numerous cell and animal studies polyphenols, including quercetin and curcumin, have been implicated in the reversal of various markers of immunosenescence (Sharma et al., 2020; Santos et al., 2021).

Polyphenols are a large family of molecules naturally occurring in plants characterized by the presence of multiple phenolic hydroxyl groups providing putative antioxidant potential. Diets high in polyphenols, especially the Mediterranean diet, have been linked to a variety of positive health outcomes. These include lower rates of degenerative diseases like atherosclerosis, as well as improved metabolic function (Bahramsoltani et al., 2019; Amiot et al., 2016). One of the central mechanisms proposed to account for some of these benefits is the effect of polyphenols on immune function and, specifically, in decreasing excessive inflammation. Additionally, it has been postulated that these molecules may exert a positive influence on the gut microbiome by acting as prebiotics, which support healthy microbial communities (Sharma et al., 2020).

While a wide range of polyphenols have been studied in preclinical and clinical trials for their effects on human health, a smaller number have been implicated for potential benefit to both epigenetic age and immunological health. This group includes the molecules quercetin and fisetin. Quercetin occurs naturally in many plants, including red onions, capers, teas, and cruciferous vegetables. Fisetin is found in fruits such as strawberries, kiwis and apples, vegetables like onions and tomatoes, and nuts.

Most research on polyphenols has followed a pharmaceutical-like model, focusing on the isolated effects of individual polyphenols on specific outcomes. However, it is notable that naturally-occurring polyphenols are delivered alongside a complex mixture of minerals, vitamins, fiber and other non-caloric plant nutrients (phytochemicals). It is suggested that the benefits of polyphenols may be synergistic when consumed alongside these other plant food components (da Silveira Vasconcelos et al., 2020; Liu et al., 2003). Therefore, naturally occurring combinations of polyphenols and other phytochemicals may be more ideally suited to exert beneficial biological effects than the same molecules consumed in isolation.

### Potential mechanism of actions of polyphenols in regulating immunity through epigenetics

Polyphenols are best understood as antioxidants primarily on the merit of in vitro studies and chemical structure. It is a widespread belief that the principal health benefit of polyphenol consumption stems from this antioxidant ability. While it is now recognized that oxidative stress (and subsequent antioxidant neutralization) may directly impact epigenetic regulation, including within immune cells, additional mechanisms of polyphenol-induced cellular effects are now being characterized (Borsoi et al, 2023). Polyphenols are poorly absorbed in the human GI tract, allowing most dietary polyphenols to reach the large intestine intact, where they are acted upon by the diverse microbes of the gut microbiome (Borsoi et al, 2023). Here, polyphenols may be modified by microbial metabolism to generate new metabolites and other bioactive compounds that may impact epigenetic regulation. For example, consumption of polyphenols may induce gut microbes to increase production of short-chain fatty acids (e.g., β-hydroxybutyrate (BHB)) which are known epigenetic regulators as well as immune modulators. Polyphenols are additionally acted on by various phase I biotransformation enzymes present in the GI tract prior to absorption, as well as phase II enzymes present in enterocytes and hepatocytes (Zhang et al, 2022). Within circulation, polyphenols and metabolites can bind to immune cells. Polyphenols may modulate epigenetics by way of DNA methylation, histone modification and miRNA expression (Marangoni et al, 2023). This has been quantified by the use of epigenetic methylation analysis of immune cell composition through the use of well-respected machine learning algorithms.

### Tartary Buckwheat

Tartary buckwheat is a buckwheat cultivar used for thousands of years for its medicinal and food properties. It is known to contain phytochemicals that include the polyphenols rutin, quercetin, luteolin and hesperidin, as well as nutrients such as d-chiro-inositol, a cyclic polyol clinically studied for its effects on human metabolism (Huda et al., 2021; Dziedzic et al., 2018; Caputo et al., 2020). The polyphenol-rich supplement to be used in this trial represents a combination of potentially epigenetically- and immunologically-active polyphenols, as well as additional phytochemicals included to better mirror the suite of biologically-active phytochemicals naturally occurring in Tartary buckwheat.

Dietary interventions with Tartary buckwheat have been independently studied and shown to have positive effects on human physiology related to immune function and metabolism (Wieslander et al., 2011; Nishimura et al., 2016). The sum of this research suggests that consumption of phytonutrients found in Tartary buckwheat may positively affect human immune function and epigenetic expression, potentially through shared pathways.

## Methods and Measurements

### Ethical Approval and Study Design

The Institute of Cellular and Regenerative Medicine Institutional Review Board granted approval for all procedures involving humans in this study. This study is registered on ClinicalTrials.gov under the registration number NCT05234203

### Statistics

As this was a pilot study, sample size was estimated based on previous analyses with epigenetic studies to require a minimum of n=30. A total of 50 participants were enrolled to account for attrition and non-compliance. Populations for statistical analysis were predefined based on inclusion and exclusion criteria (see below). Subgroup and sensitivity analyses were conducted based on +1 and -1 standard deviations from the mean and within/inclusive of 1 standard deviation of the mean and those testing positive to COVID-19 during the study, as accelerated epigenetic aging in the context of COVID-19 has been documented (Cao et al., 2022). Demographics and analysis of the GHQ were conducted in Excel (Microsoft, version 2211) and presented as descriptive statistics.

### Participant Recruitment and Eligibility

50 generally healthy men and women between the ages of 18 and 85 years (inclusive) with body mass index (BMI) <40 kg/m^2^ were enrolled in the study. Participants were required to have an established primary care provider and active health insurance; be able to read, write, and speak English fluently; and be able to comply with the protocol instructions including performing the in-home venous blood draw using a Tasso device. Investors or immediate family members possessing investment in Big Bold Health were excluded from the study.

Women who were pregnant and/or lactating and individuals on jobs requiring night shift work were excluded. Exclusion criteria also included history (prior 2 years) or presence of cancer, except for non-melanoma skin cancer; known history of blood dyscrasias including coagulopathy or use of prescription anticoagulants; diagnosis of a transient ischemic attack (within 6 months); presence of clinically significant acute or unstable cardiovascular or cerebrovascular disease, psychiatric disorder, alcohol or chemical dependence; immune-related conditions (e.g., hepatitis C, HIV, or active infection within the previous 4 weeks) or other illness that in the opinion of the Clinical Investigator would render a participant unsuitable to participate in the study. In addition, those with known allergy to any of the components of the test product, those consuming known prescription immunomodulating products (e.g. oral glucocorticoids, TNF-α inhibitors) or concentrated polyphenolic supplements within 1 month the baseline visit. Concentrated polyphenolic substances that were specifically excluded prior to, and during the study were quercetin, rutin, luteolin, epigallocatechin gallate (EGCG), resveratrol, curcumin, fisetin, berberine, soy isoflavones (genistein, daidzein, and glycitein), hesperidin, and ellagic acid.

### Study Objectives

The primary objective of this exploratory clinical trial was to evaluate the effect of consuming a polyphenol-rich supplement largely based around the phytochemical composition of Tartary buckwheat (HTB Rejuvenate) for 90 days on measurements of epigenetically-measured immune age.

The secondary objective was to assess the effects of HTB Rejuvenate on peripheral leukocyte immune profiles after 90 days, as well as on GO pathways. Tertiary objectives included clinical observations using a General Health Questionnaire (GHQ). Safety was also assessed via reports of AEs.

### Study Product

The study product was a polyphenol-rich supplement (HTB Rejuvenate) that is commercially available and produced under Good Manufacturing Practices (GMP). The HTB Rejuvenate supplement delivered 579 mg of polyphenols per serving (2 capsules), which is within the studied ranges of polyphenol intake consumed through diet (Del Bo et. al., 2019).

The study product is a concentrated version of naturally occurring phytonutrients in Tartary buckwheat that contains rutin, quercetin, luteolin, hesperidin as well as d-chiro-inositol (**TABLE 1**). 2-hydroxybenzylamine (2-HOBA) is an antioxidant phytochemical also isolated from Tartary buckwheat. Hydroxymethylbutyrate (HMB) was also included in the study product as this is a naturally-occuring byproduct of leucine metabolism linked to improvement in age-related metrics including muscle loss (Engelen and Deutz, 2018). Tartary buckwheat is particularly enriched in leucine (Dong et al., 2023). Test product was consumed at 4 capsules per day, taken as 2 capsules twice per day close to 12 hours apart. Specifically, participants were counseled to consume 2 capsules in the morning (between 6 and 10 am) and 2 capsules in the evening (between 6 and 10 pm) with food.

**TABLE 1:** Composition of HTB Rejuvenate.

| Component | Amount per Serving<br>(2 capsules) | Amount per day<br>(4 capsules) |
| --- | --- | --- |
| Himalayan Tartary buckwheat (HTB) flour | 95 mg | 190 mg |
| D-chiro inositol (DCI/D-chiro-inositol) | 150 mg | 300 mg |
| 2-hydroxybenzylamine (2-HOBA) | 13 mg | 26 mg |
| Hydroxymethylbutyrate (HMB) | 69 mg | 138 mg |
| Chlorophyllin | 7.5 mg | 15 mg |
| Polyphenols* | 579 mg | 1,158 mg |
| Vitamin C | 20 mg | 40 mg |
| Calcium | 30 mg | 60 mg |

\* Polyphenols delivered in one serving (2 capsules) included 330 mg quercetin, 83 mg rutin, 83 mg hesperidin, and 83 mg luteolin

### Analysis Populations

A review of compliance and protocol deviations was conducted prior to data analysis, and those participants whose data is included for analysis populations is summarized in **Appendix 1**. Based on the completion of the blood sampling for the epigenetic tests, the modified intent to treat (ITT) population, which includes all participants who completed both baseline and final labs, was composed of n=47. Two participants who did not complete the final blood draw were excluded from the ITT population because the outcome required both tests for analysis. In addition to removal of the two individuals who were excluded from the ITT, the per-protocol (PP) population excluded seven other participants, including two for use of excluded medications/supplements, four for low study supplement compliance, and one for both inclusion of an excluded medication/supplement and low compliance.

### Demographics

The demographics obtained during screening/baseline clinical interviews for the ITT and PP populations are provided in **TABLE 2**. Participants in the PP population, which was the primary population for the laboratory analyses, were 54 y (SD, 11 y) old with BMI of 24.2 kg/m^2^ (SD, 3.3 kg/m^2^). A total of 14 participants had documented cases of COVID during the study (See **Appendix 1**).

**TABLE 2:** Characteristics of Participants.

| Characteristic | Units | ITT | PP |
| --- | --- | --- | --- |
| Population Number | N | 47 | 40 |
| Age | y (±SD) | 54 (±11) ‡ | 54 (±11) § |
| Sex | % Female | 60 | 62.5 |
| Weight (self-report)† | kg | 70.3 (±15.7) ‡ | 69.2 (±14.7) § |
| Height (self-report)† | cm | 166.7 (±11.1) ‡ | 166.5 (±11.2) § |
| BMI | kg/m<sup>2</sup> | 24.6 (±3.5) | 24.2 (±3.3) |

Abbreviations: BMI, body mass index; cm, centimeter; ITT, intent-to-treat; kg, kilogram; m, meter; N, subject number; PP, per protocol; SD, standard deviation; y, years
\*The ITT data excluded the two participants who did not complete final lab tests.
†Self-reported weight and height data were converted to metric units for reporting purposes.
‡The ITT population had no data for 3 participants for age, and 11 participants for weight and height. All participants had reported BMI data.
§The PP population had no data for 3 participants for age, and 7 participants for weight and height. All participants had reported BMI data.

### Study Design

This was a virtual, single-arm open-label prospective pilot clinical trial comparing epigenetic and immune assays in participants prior to and after 90 days of supplementation with the polyphenol-rich HTB Rejuvenate supplement. The study included a screening/baseline visit (Visit 1, Day -15), communication for starting product (Visit 2, Day 0), 24-hour check-in after product start (Visit 3, Day 1), and three follow-up visits (Visit 4, Day 30. Visit 5, Day 60; Visit 6, Day 90). In addition, participants were contacted by phone approximately 30 to 90 d after completing the study and when laboratory results were available. Study details are provided in the Study Tracking Table (**TABLE 3**).

**TABLE 3:**
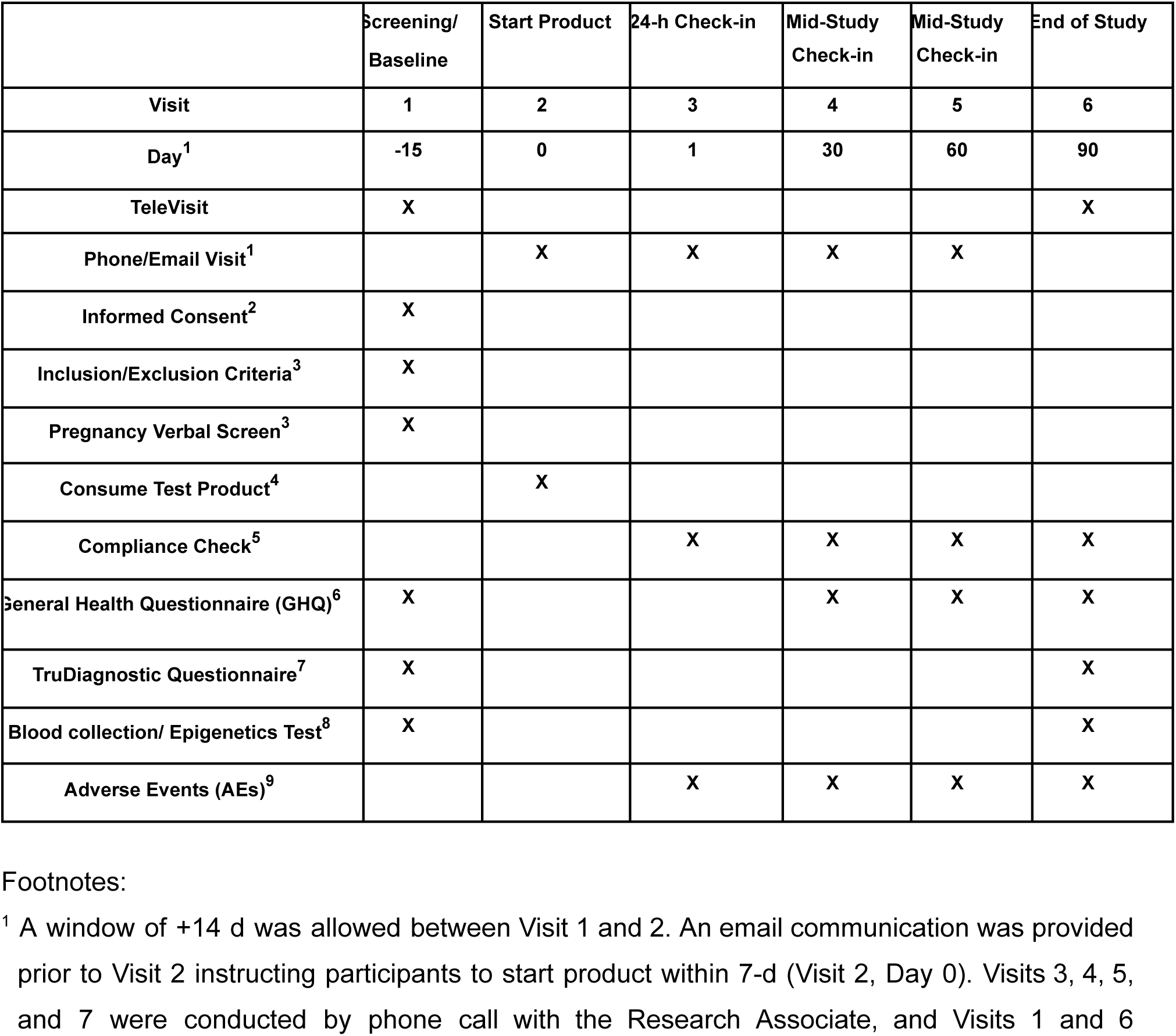
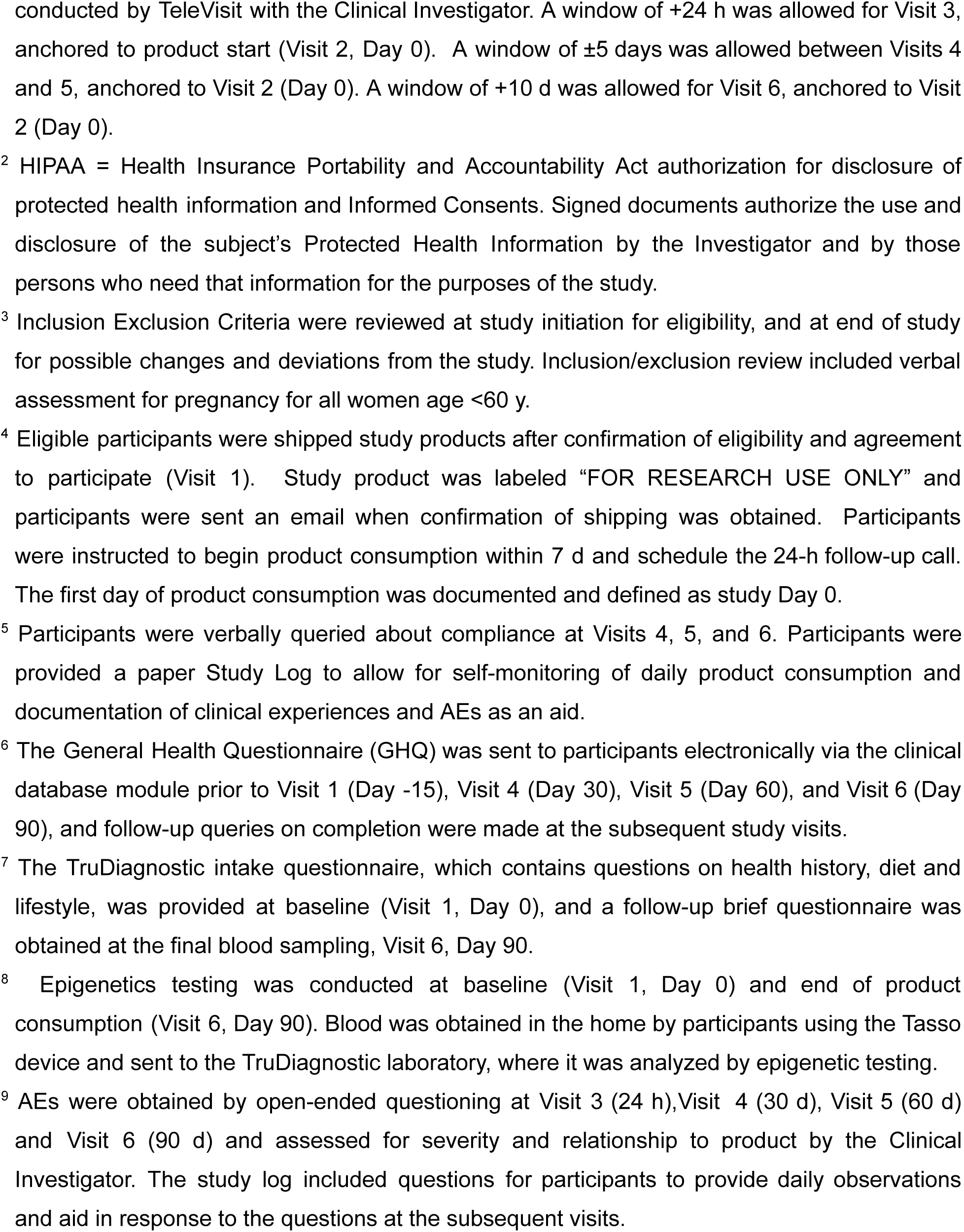
Study Tracking Table.

|  | Screening/<br>Baseline | Start Product | 24-h Check-in | Mid-Study<br>Check-in | Mid-Study<br>Check-in | End of Study |
| --- | --- | --- | --- | --- | --- | --- |
| Visit | 1 | 2 | 3 | 4 | 5 | 6 |
| Day<sup>1</sup> | -15 | 0 | 1 | 30 | 60 | 90 |
| TeleVisit | X |  |  |  |  | X |
| Phone/Email Visit<sup>1</sup> |  | X | X | X | X |  |
| Informed Consent<sup>2</sup> | X |  |  |  |  |  |
| Inclusion/Exclusion Criteria<sup>3</sup> | X |  |  |  |  |  |
| Pregnancy Verbal Screen<sup>3</sup> | X |  |  |  |  |  |
| Consume Test Product<sup>4</sup> |  | X |  |  |  |  |
| Compliance Check<sup>5</sup> |  |  | X | X | X | X |
| General Health Questionnaire (GHQ)<sup>6</sup> | X |  |  | X | X | X |
| TruDiagnostic Questionnaire<sup>7</sup> | X |  |  |  |  | X |
| Blood collection/ Epigenetics Test<sup>8</sup> | X |  |  |  |  | X |
| Adverse Events (AEs)<sup>9</sup> |  |  | X | X | X | X |

<sup>1</sup> A window of +14 d was allowed between Visit 1 and 2. An email communication was provided prior to Visit 2 instructing participants to start product within 7-d (Visit 2, Day 0). Visits 3, 4, 5, and 7 were conducted by phone call with the Research Associate, and Visits 1 and 6
conducted by TeleVisit with the Clinical Investigator. A window of +24 h was allowed for Visit 3, anchored to product start (Visit 2, Day 0). A window of $\pm 5$ days was allowed between Visits 4 and 5, anchored to Visit 2 (Day 0). A window of +10 d was allowed for Visit 6, anchored to Visit 2 (Day 0).
<sup>2</sup> HIPAA = Health Insurance Portability and Accountability Act authorization for disclosure of protected health information and Informed Consents. Signed documents authorize the use and disclosure of the subject's Protected Health Information by the Investigator and by those persons who need that information for the purposes of the study.
<sup>3</sup> Inclusion Exclusion Criteria were reviewed at study initiation for eligibility, and at end of study for possible changes and deviations from the study. Inclusion/exclusion review included verbal assessment for pregnancy for all women age <60 y.
<sup>4</sup> Eligible participants were shipped study products after confirmation of eligibility and agreement to participate (Visit 1). Study product was labeled "FOR RESEARCH USE ONLY" and participants were sent an email when confirmation of shipping was obtained. Participants were instructed to begin product consumption within 7 d and schedule the 24-h follow-up call. The first day of product consumption was documented and defined as study Day 0.
<sup>5</sup> Participants were verbally queried about compliance at Visits 4, 5, and 6. Participants were provided a paper Study Log to allow for self-monitoring of daily product consumption and documentation of clinical experiences and AEs as an aid.
<sup>6</sup> The General Health Questionnaire (GHQ) was sent to participants electronically via the clinical database module prior to Visit 1 (Day -15), Visit 4 (Day 30), Visit 5 (Day 60), and Visit 6 (Day 90), and follow-up queries on completion were made at the subsequent study visits.
<sup>7</sup> The TruDiagnostic intake questionnaire, which contains questions on health history, diet and lifestyle, was provided at baseline (Visit 1, Day 0), and a follow-up brief questionnaire was obtained at the final blood sampling, Visit 6, Day 90.
<sup>8</sup> Epigenetics testing was conducted at baseline (Visit 1, Day 0) and end of product consumption (Visit 6, Day 90). Blood was obtained in the home by participants using the Tasso device and sent to the TruDiagnostic laboratory, where it was analyzed by epigenetic testing.

**Table 4:** Investigation of epigenetic age measures based on subsets one standard deviation higher than the mean, one standard deviation lower than the mean, and within or equal to one standard deviation (-1 to +1) of the mean for multiple epigenetic aging algorithms.

|  | Mean -<br>Test 1 | SD -<br>Test 1 | Mean -<br>Test 2 | Mean - SD<br>Test 2 | N | Wilcoxon<br>(p-value) |
| --- | --- | --- | --- | --- | --- | --- |
| OMICmAge EAA - 1SD Higher | 5.686 | 1.176 | 4.185 | 2.904 | 7 | 0.380 |
| OMICmAge - 1SD Higher | 61.290 | 10.649 | 59.940 | 11.844 | 7 | --- |
| **OMICmAge EAA - 1SD Lower** | **-4.969** | **1.037** | **-3.718** | **1.851** | **7** | **0.031** |
| OMICmAge - 1SD Lower | 48.032 | 5.557 | 49.404 | 5.709 | 7 | --- |
| OMICmAge EAA - Within 1SD | -0.291 | 1.906 | -0.031 | 3.120 | 26 | 0.860 |
| OMICmAge - Within 1SD | 54.785 | 7.201 | 55.268 | 7.538 | 26 | --- |
| OMICmAge EAA - All | -0.063 | 3.604 | 0.062 | 3.699 | 40 | 0.740 |
| OMICmAge - All | 54.741 | 8.439 | 55.059 | 8.548 | 40 | --- |
| **PCPhenoAge EAA - 1SD Higher** | **8.065** | **2.163** | **4.262** | **2.294** | **6** | **0.031** |
| PCPhenoAge - 1SD Higher | 51.004 | 4.785 | 47.456 | 6.345 | 6 | --- |
| PCPhenoAge EAA - 1SD Lower | -9.300 | 2.886 | -5.790 | 4.221 | 7 | 0.078 |
| PCPhenoAge - 1SD Lower | 36.754 | 11.490 | 40.537 | 11.347 | 7 | --- |
| PCPhenoAge EAA - Within 1SD | -0.087 | 2.435 | 0.166 | 4.132 | 27 | 0.360 |
| PCPhenoAge - Within 1SD | 48.759 | 10.207 | 49.251 | 10.601 | 27 | --- |
| PCPhenoAge EAA - All | -0.476 | 5.580 | -0.262 | 4.854 | 40 | 0.690 |
| PCPhenoAge - All | 46.995 | 10.778 | 47.457 | 10.522 | 40 | --- |
| PCGrimAge EAA - 1SD Higher | 2.960 | 0.822 | 2.177 | 1.455 | 8 | 0.200 |
| PCGrimAge - 1SD Higher | 63.209 | 11.339 | 62.561 | 11.717 | 8 | --- |
| ***PCGrimAge EAA - 1SD Lower*** | **-3.759** | **0.554** | **-1.333** | **0.536** | **6** | **0.031** |
| PCGrimAge - 1SD Lower | 57.527 | 12.238 | 60.078 | 12.357 | 6 | --- |
| PCGrimAge EAA - Within 1SD | -0.407 | 1.165 | -0.336 | 2.166 | 26 | 0.670 |
| PCGrimAge - Within 1SD | 61.605 | 7.425 | 61.855 | 7.860 | 26 | --- |
| PCGrimAge EAA - All | -0.236 | 2.248 | 0.017 | 2.178 | 40 | 0.320 |
| PCGrimAge - All | 61.314 | 8.979 | 61.730 | 9.187 | 40 | --- |
| PACE - 1SD Higher | 1.007 | 0.050 | 0.976 | 0.071 | 8 | 0.250 |
| PACE - 1SD Lower | 0.710 | 0.033 | 0.699 | 0.090 | 7 | 0.690 |
| PACE - Within 1SD | 0.841 | 0.050 | 0.866 | 0.079 | 25 | 0.110 |
| PACE - All | 0.851 | 0.104 | 0.859 | 0.116 | 40 | 0.610 |

Participants were pre-screened using an on-line questionnaire and those meeting criteria were provided a detailed study description and the electronic Informed Consent. Interested individuals attended a TeleVisit (Visit 1, Day -15) with the Clinical Investigator, during which it was confirmed that they met all of the inclusion and none of the exclusion criteria. Eligible participants who voluntarily signed the Informed Consent were then provided the baseline GHQ electronically and sent the Tasso blood draw device and instructions to obtain an in-home blood sample for the baseline epigenetic testing. Participants were also sent two bottles of study product (120 capsules/bottle) and instructions for starting consumption after baseline data were obtained (Visit 2, Day 0), and scheduling the 24-h call (Visit 3, Day 1). Participants were instructed not to make substantial changes to diet and lifestyle (e.g., dietary and physical activity programs, new supplements) during study period unless explicitly recommended by their treating medical provider.

At Visit 3, participants were asked about adverse events (AEs), product consumption start date was confirmed, and participants were reminded of the study instructions, including not consuming polyphenol supplements and maintaining habitual diet and lifestyle during the study. Mid-study check-in visits were conducted by phone call with the Research Associate at Visit 4 (Day 30) and Visit 5 (Day 60), during which participants were queried for compliance, AEs, and changes to concomitant medications/supplements and habitual diet and lifestyle. In addition, participants were sent the GHQ electronically and queried on completing the questionnaire. Bottles of study product (30 capsules) were sent after Visit 4 and Visit 5.

Prior to the end-of-study visit (Visit 6, Day 90), participants were sent the link to the final GHQ as well as a Tasso blood draw kit. At Visit 6, participants attended a TeleVisit with the Clinical Investigator for the final review, which included study product consumption compliance, completion of the final GHQ, AEs, maintenance of habitual diet and lifestyle, and any changes to concomitant medications and supplements. Confirmation that the final blood was obtained before participants discontinued the study product.

### Clinical and Anthropometric Information

Relevant medical history were reviewed by the Clinical Investigator or appropriate designee prior to signing the Informed Consent. Anthropometric information included self-reported height and weight, with the date when the participant last weighed themselves documented, and BMI was calculated. Participants were questioned on medication and supplement use at Visit 1 (Day -15) and asked about changes to medication and supplement use, as well as diet and lifestyle at Visits 4, 5, and 6 (Days 30, 60, and 90).

### General Health Questionnaire (GHQ)

Participants were asked to complete an electronic GHQ (**Appendix 2**) at baseline and 30-d, 60-d, and 90-d after the start of test product consumption. The GHQ included questions on general health over the previous 4 weeks with answers scored on a 5-point balanced Likert scale. The Likert scale varied randomly from lowest to highest score for questions 3, 4, 7, 9, 11, 12, 13, and 15, and from highest to lowest score for questions 1, 2, 5, 6, 8, 10, and 14 to decrease order-effects. In addition, leading and subjective language were avoided in the question design. The individual health categories included: 1: General Health: questions 1, 2, and 13, score range 3 to 15 points. 2: Gastrointestinal (GI): questions 3 and 4, score range 2 to 10 points. 3: Energy & mood: questions 5, 10, 11, 12, 14, and 15, score range 6 to 30 points. 4: Allergy & Skin: questions 6, 7, 8, and 9, score range 4 to 20 points.

### Compliance

Participants were provided a paper study log with the initial product shipment that included a tracking form for documenting the consumption of 2 capsules of study product in the morning and 2 capsules in the evening every day, as well as a section for daily notes for unusual symptoms, illness and reasons for missed product consumption. The study log was not collected but provided to aid participants in answering queries at the relevant visits. Compliance was obtained during Visits 4, 5, and 6 by query and documented as % consumed over the previous 4 weeks from data on the study logs, as verbally provided by participants.

### Epigenetic Age Biomarkers

The standardized epigenetic biological age test examined methylation at 850,000 DNA locations on CpG sites and was conducted by TruDiagnostic at baseline and after 90 days of study product consumption. Blood samples were obtained by participants using the in-home Tasso device and shipped to the TruDiagnostic laboratory directly. Overall, we analyzed blood samples from 47 adults from an initial sample set of 50 individuals using an Illumina epigenetic panel that analyzed DNA methylation at 850,000 CpG sites prior to and after 90 days of an intervention with a polyphenol-rich supplement designed to mimic major bioactive nutrients found in the plant Tartary buckwheat. Beta values were extracted from IDAT files using the minfi pipeline, and outlier samples were identified using the ENmix R package (Aryee et al., 2014; Xu et al. 2016). Final analysis included data from 40 people who successfully completed the study. All analyses were conducted using the R programming environment.

Epigenetic clocks were derived from processed beta values included the Horvath multi-tissue clock (Horvath 2013), the Horvath skin and blood clock (Horvath et al., 2018), the Hannum clock (Hannum et al,. 2013), the PhenoAge clock (Levine et al. 2018), GrimAge versions 1 (Lu et al., 2019), and the DNAmTL clock (Lu, Seeboth, et al. 2019). To ensure high reproducibility, the principal component versions of these clocks were used as outlined by Higgins-Chen et al. (Higgins-Chen et al. 2022). Individual system clocks were calculated following the methodology described by Sehgal et al. (Sehgal et al. 2023). The clocks were calculated using a custom R script available on GitHub. Additionally, DunedinPACE was computed using a specific script available on GitHub (https://github.com/danbelsky/DunedinPACE, (Belsky et al. 2022)). Epigenetic assessment was conducted using Wilcoxon-rank sum test performed by evaluators blinded to participant identifying information. For all statistical tests, epigenetic measures were converted from raw outputs to epigenetic age acceleration (EAA), which is defined as the residual calculated between the epigenetic age measure and chronological age. To account for batch effects, the variation calculated from the first three principal components of the control probes were added as fixed effects to the EAA estimation. Epigenetic assessment was conducted using Wilcoxon-rank sum test performed by evaluators blinded to participant identifying information.

Immune age assessments were obtained via deconvolution algorithms of the epigenetic data, which quantitatively approximate immune cell subsets including CD4+ T cells, CD8+ T cells, granulocytes, natural killer (NK) cells, monocytes, eosinophils, and neutrophils using the Epidish package (Luo et al., 2023).

### Differential Methylation Analysis

Differential methylation analysis was carried out utilizing processed beta values. The Limma package was employed for three distinct comparisons, identifying differentially methylated loci (DMLs) within the individuals during the trial from baseline to the final timepoint collection (day 90). Multivariate linear models incorporated fixed effects including beadchip, five immune cell percentages (CD4T naive, CD4T memory, CD8T naive, CD8T memory, B naive, B memory, Eosinophils, T regulatory, NK, and Neu), the third and fourth three principal components of technical probes, and the Study ID for the individual. Q-Q plots and lambda values were utilized to assess P-value inflation or deflation across the methylome (Guintivano et al., 2020); the final lambda value of was 0.96 suggesting the EWAS model did not show inflation or deflation. DMLs were identified with an unadjusted p-value significance threshold of < 0.001. Functional annotation of DMLs was conducted using the GREAT pipeline to identify significant gene ontology terms, as implemented in the rGREAT R package (Gu and Hübschmann 2023).

### Safety Assessments

Participants were asked about AEs by open-ended questions at Visit 3 (24-h), Visit 4 (Day 30), Visit 5 (Day 60), and Visit 6 (Day 90). The Clinical Investigator reviewed the reports, obtaining follow-up information from participants as needed, and categorized by severity and relationship to study products based upon FDA criteria (detailed in the Study Protocol).

## Results

### Participant Disposition

Fifty-seven individuals were screened on-line and scheduled for an initial visit to assess eligibility. Of these, n=50 individuals attended the on-line screening visit and met eligibility criteria, and n=7 did not meet eligibility criteria, as follows:

- Involved in another trial, n=2
- Lactating, n=1
- On excluded high dose polyphenolic supplements, n=1
- BMI outside accepted ranges, n=1
- Did not show up for screening, n=1
- Did not want name associated with the DNA results in the database n=1

Of the n=50 enrolled in the study, all but one completed the visits. One participant withdrew consent at Visit 2. In addition, final lab analyses were not obtained from two participants (n=2) and these were considered early terminations (ET) at Visit 6 for purposes of analyses. The participant disposition is summarized in **Figure 1**.

**Figure 1:**
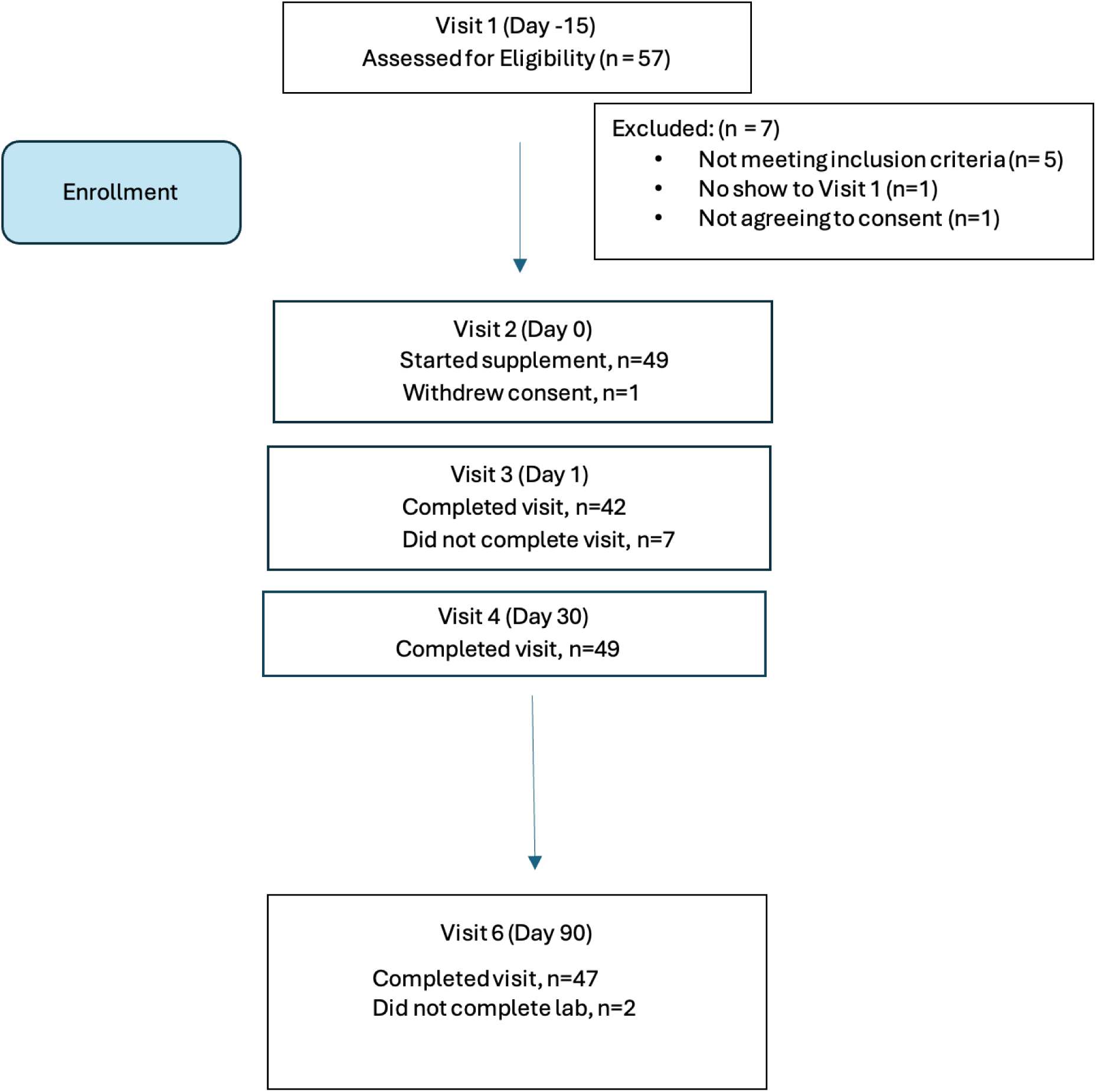
Participant Disposition Chart

**Figure 2:**
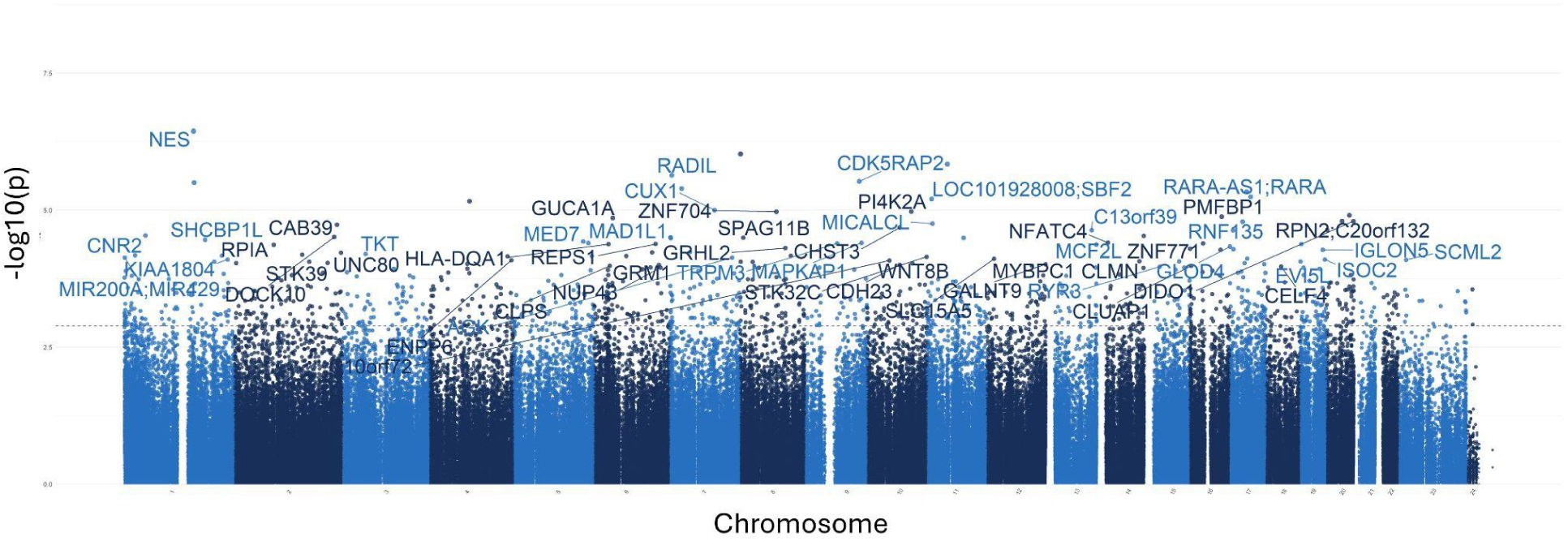
Manhattan plots for the epigenome-wide association study (EWAS)

### EWAS analysis identifies CpGs related with intake of Tartary buckwheat extract

To investigate the overall epigenetic impact of the standardized polyphenol concentrate upon the cohort, we conducted an epigenome-wide analysis (EWAS) analysis to identify CpGs which showed significant differential methylation between the two visits. To ensure that the EWAS model was not overfit and thus limit false positives, we first tested for inflation by identifying variables which accounted for overfitting. To this end, we estimated the coefficient of overfitting (lambda) at 0.96, which indicated that the EWAS model selected did not show significant overfitting within the data (Supplementary File X). From the indicated model, we identified 887 Differentially Methylated Loci (DMLs) across the EPIC/850K data (unadjusted p-value < 0.001). Among these, 336 CpG sites showed hypermethylation at the conclusion of the study (Visit 6), while 551 loci were hypomethylated at the conclusion of the study. The full list of CpGs is listed in **Supplementary File 2** for the analysis.

The above plot depicts genes associated with CpG sites identified in the analysis. Each dot on the plot represents a CpG site, with its vertical position corresponding to the negative logarithm (base 10) of the unadjusted p-value for DNA methylation association, with a significance threshold set at p = 0.001. The x-axis shows genomic positions organized by chromosomes, with color-coded dots indicating specific chromosomes. The prominently peaked dots represent CpG sites that surpass the genome-wide significance threshold, indicating significant associations.

An epigenome-wide association study and enrichment analysis was conducted to compare pre- and post-intervention data (90 days after starting the study supplement). The Volcano plot illustrates differentially methylated loci (DMLs) identified in the pre- vs post-intervention comparison. Each dot represents a CpG site, with its vertical position indicating the negative logarithm (base 10) of the unadjusted p-value for DNA methylation association. The x-axis shows the relative log fold change (logFC) of the m-values between the two timepoints. Negative values indicate CpGs with decreased methylation among study participants (green), while positive values indicate increased methylation (red).

### Gene Ontology Pathways

To link the methylation results to biological processes, enrichment analyses using the GREAT software were conducted on CpGs based on the direction of methylation, and used to identify gene ontology (GO) pathways. Hypermethylated CpGs at the conclusion of the study were significantly associated with a total of 15 GO-BP (Biological processes) terms, 4 GO-MF (molecular function), and 3 GO-CC (Cellular component) (**Supplementary File 3**) terms. The top 15 terms for each GO category is shown in **Figure 4** below, which included enrichment of GO-BP terms involved in DNA repair (*regulation of double-strand break*), development (*labyrinthine layer morphogenesis, neural crest migration, embryonic placenta development*) as well as ceramide kinase activity and *COP9 signalosome activity*.

**Figure 3:**
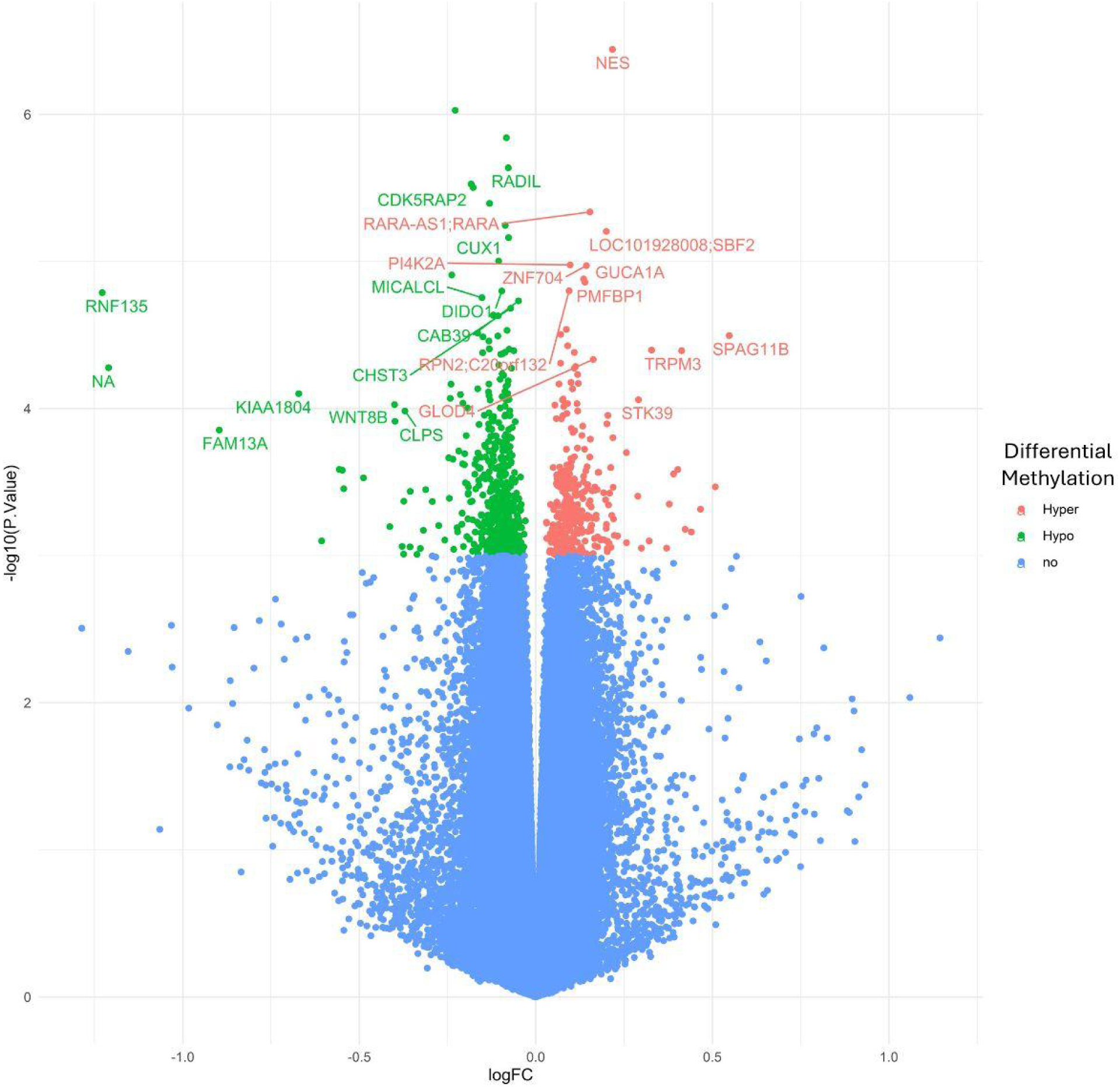
Volcano plot for epigenome-wide association study and enrichment analysis

**Figure 4:**
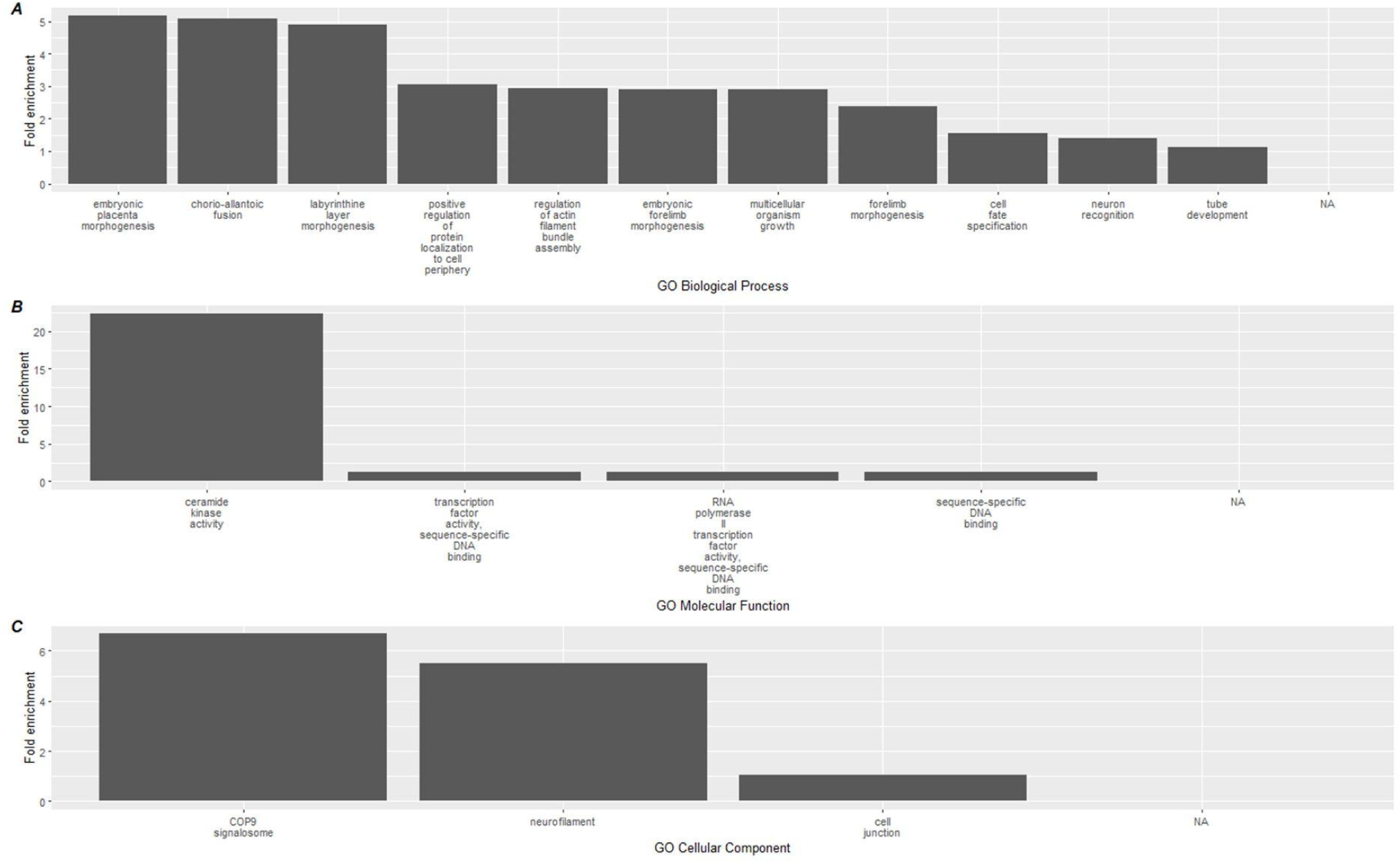
Top 15 significant gene ontology terms associated with hypermethylated DMLs using GREAT.

Gene ontology databases are reported for A) GO-BP, B) GO-MF, and C) GO-CC. Biological associations shown are geneontology (GO) terms for Biological Processes (BP), Molecular Function (MF), and cellular components (CC).

A similar analysis was performed on the hypomethylated DMLs identified in the analysis, which revealed greater significant GO terms compared to the hypermethylated DMLs (**Supplementary File 3**). Among the hypomethylated DMLs, 124 GO-BP terms, 6 GO-MF terms, and 4 GO-CC terms. The top 15 for each category are reported in **Figure 5** which includes the activation of processes that negatively regulate photoreceptor cell differentiation, and increase positive regulation of fibroblast proliferation. In addition, we also observed higher enrichment of processes associated with Notch binding.

**Figure 5:**
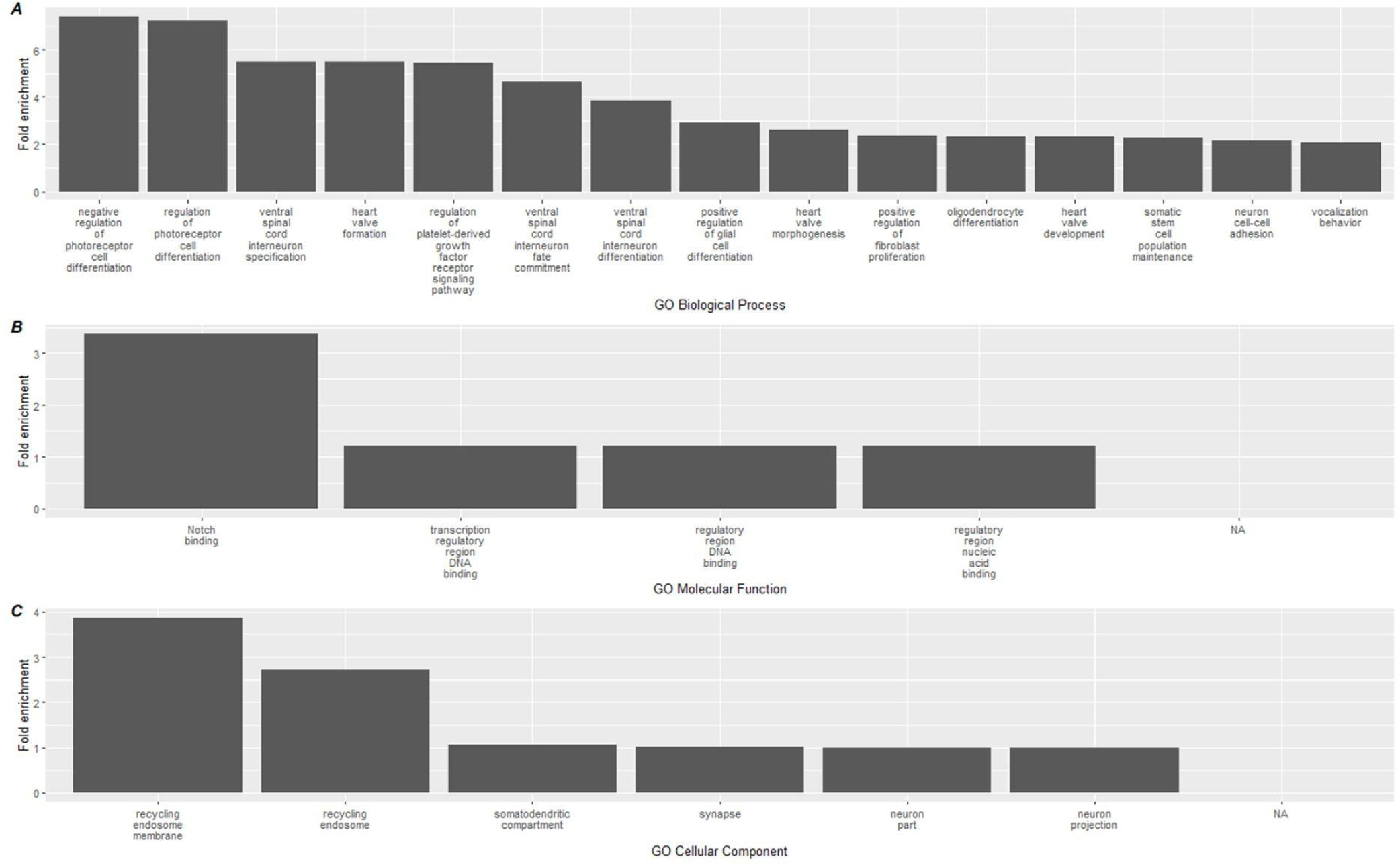
Top 15 significant gene ontology terms associated with hypomethylated DMLs using GREAT. Gene ontology databases are reported for A) GO-BP, B) GO-MF, and C) GO-CC.

**Figure 6:**
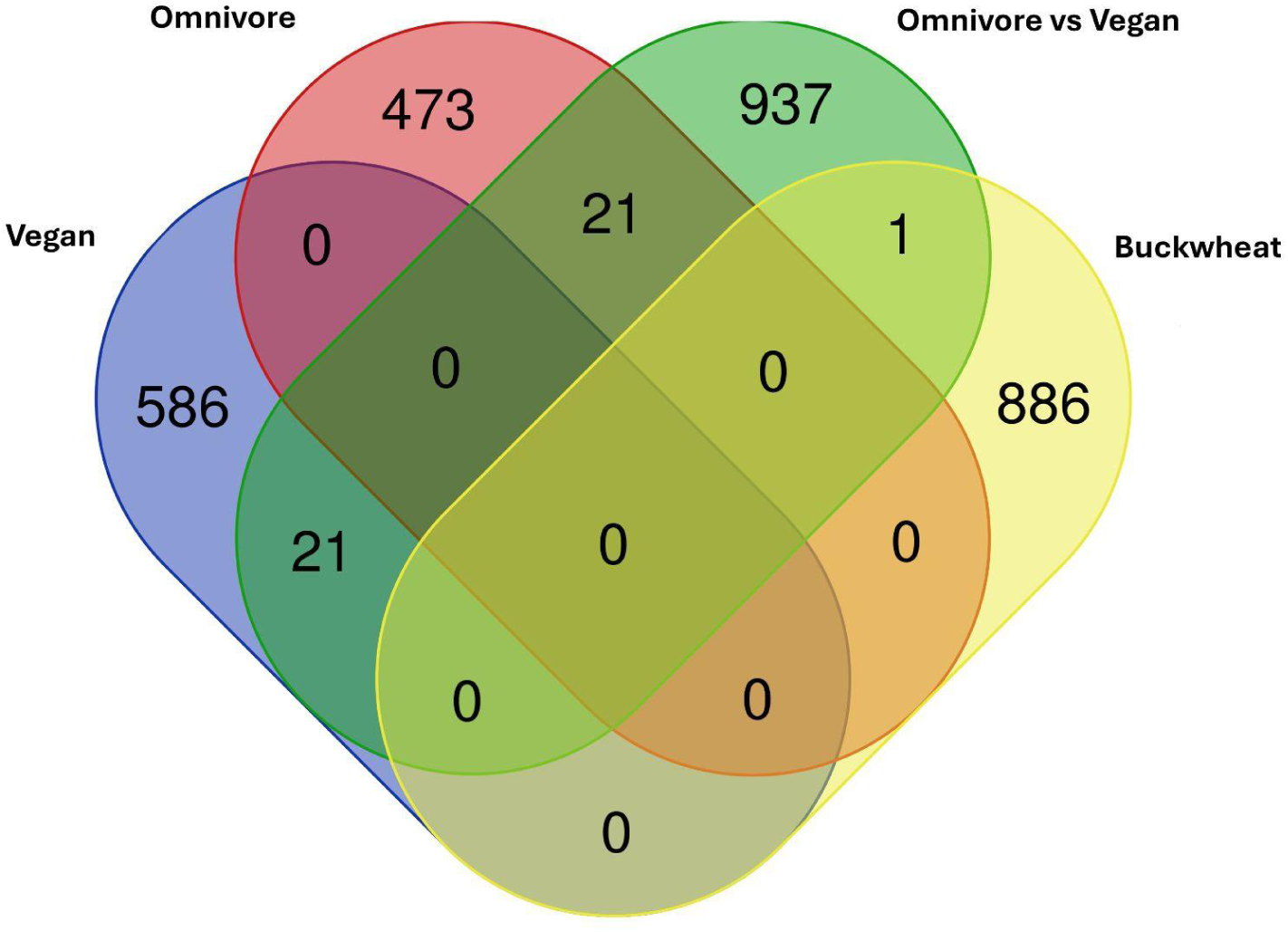
Venn Diagram of DMLs identified between the comparisons in Dwaraka et al. 2024 plotted against DMLs from the Tartary buckwheat supplement intervention.

### Comparison to Published Dietary DML Data

To better interrogate whether the above results might represent changes seen in more comprehensive dietary intervention, and whether the study supplement mimicked the effects of a plant-based diet, we compared our data to the results from Dwaraka et al., 2024, which examined DMLs across a vegan intervention and omnivore (control) intervention using a Venn diagram of DMLs across the three groups.

The one CpG shared among the current analysis with the Omnivore analysis is cg05093714 (Gene ID: LINC01095) which is significantly higher in the vegan cohort compared to the omnivore cohort at 8 weeks. However, all other CpGs are specific to the Tartary buckwheat cohort identified here. No overlap is observed between the Vegan or Omnivore diet, suggesting different pathways are involved.

### Study supplement use impacts changes in epigenetic age

In order to determine the response to the study supplement on biological age, we quantified and performed analysis on a host of biological age metrics using DNA methylation. Aging clocks used included the second generation multi-omic informed OMICmAge, the third generation DunedinPACE (PACE) and principal component (PC) based second generation PhenoAge and GrimAge clocks. We additionally utilized epigenetic age acceleration (EAA), a marker of the difference between expected rate of aging based on chronological and biological aging. Remarkable findings included: 1. A slowing of epigenetic age acceleration in people with a PCPhenoAge 1SD Higher than the mean (p = 0.031) 2. An increase in epigenetic age acceleration in people with a PCGrimAge 1SD Lower than the mean (p = 0.031) 3. An increase in epigenetic age acceleration in people with a OMICmAge 1SD Lower than the mean (p = 0.031). Note that similar p values are a result of a wilcoxon-rank sum test which utilizes a rank based estimate used against smaller n values. We stratified sample groups by subsets one standard deviation higher than the mean, one standard deviation lower than the mean, and within/including one standard deviation (-1 to +1) of the mean to better understand the degree to which starting epigenetic state impacts outcome, and as epigenetic changes seen in the context of dietary modification are typically more subtle.

It is important to note the heterogeneity of individual results measured using age-related algorithms across the 90-day study period. Individual slopes calculated as changes in signify changes in epigenetic age metrics during this window (positive slopes represent increased epigenetic age acceleration, negative slopes represent decreased epigenetic age acceleration). These results may speak to individual differences in epigenetic sensitivity to environmental inputs, in this case dietary modification. A representative analysis of changes in EAA in subset populations is shown below in **Figure 7**. A representative analysis of individual epigenetic age score changes is shown below in **Figure 8**.

**Figure 7:**
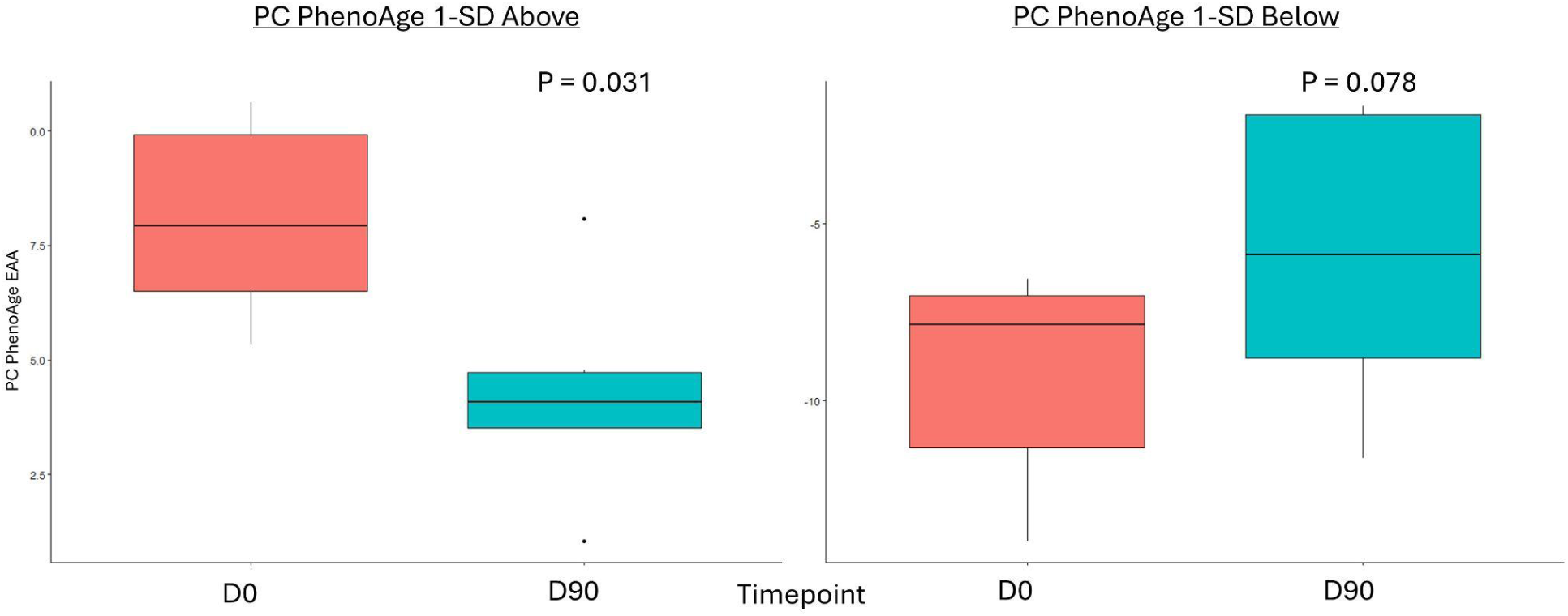
Boxplot showing the changes in PCPhenoAge EAA calculated from the subset of participants starting with a PCPhenoAge EAA one SD above the mean (red box on left compared to teal box on left) and those starting with a PCPhenoAge EAA one SD below the mean (red box on right compared to teal box on right).

**Figure 8:**
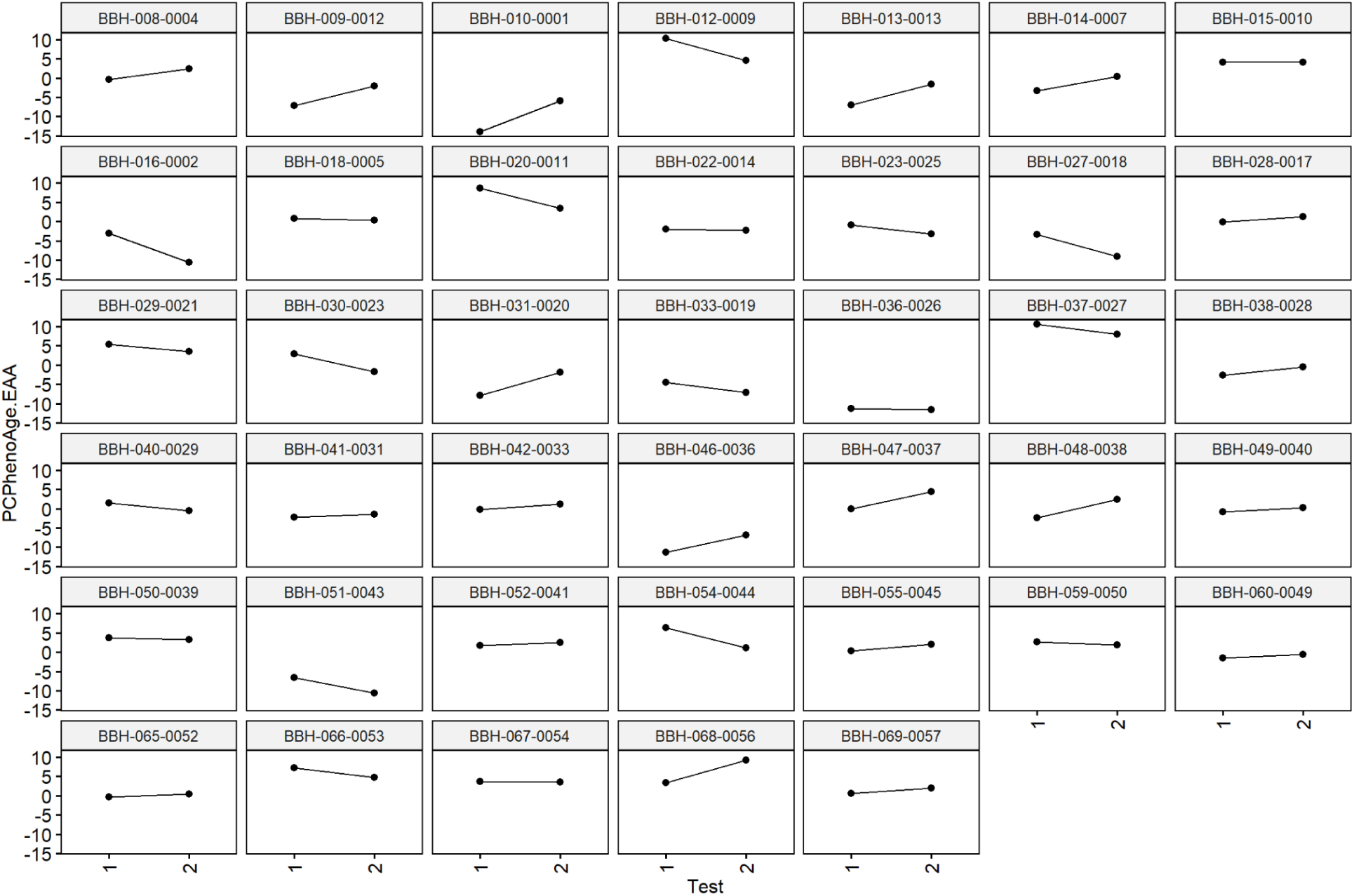
Visual representation of individual epigenetic age accelerations (PCPhenoAge used as example). Each graph is faceted by each individual’s de-identified ID provided during the trial.

### Deconvoluted Immune Cell Analysis

Alterations in immune cell makeup and function have been studied in references to both dietary change and specific nutrient augmentation. We used deconvolution methods to determine immune cell population changes over the duration of the study period to explore the effects of study supplementation on immune cell parameters across different biological age algorithms and within subsets of study participants.

### Analysis of GHQ results

The GHQ data were reviewed for completeness prior to analysis. Results were obtained for all 49 participants for the baseline and 90-d questionnaires. However, 10 datapoints were missing from the 30-d questionnaires and five datapoints were missing in the 60-d data set. Given this was the first virtual study with this questionnaire, learnings on how easy it was to obtain the data, as well as whether participants were consistent in reporting on the questionnaire was of use. As shown in **TABLE 6**, the time between questionnaires ranged widely.

Two participants indicated taking the baseline GHQ after the start of supplement, which meant the true baselines were not available for these individuals. Therefore, their data were removed from further analysis of the GHQ. In addition, two participants began supplements that could directly affect the data and were also removed. Therefore, a total of N=36 responses were reviewed for the GHQ. Summary statistics from these respondents are provided in **Appendix 3**.

In general, the population began the study in very good health by self-report. Overall, few changes were noted in any category, however, the population began with middle to high ratings in all areas of health. Therefore, changes in GHQ-measured health scores were not likely to be observed in this population.

## Discussion

Dietary interventions for modulating health have been well-documented for millenia. In the last hundred years, we’ve increasingly understood that food represents a complex mixture of caloric and non-caloric components capable of impacting physiology through myriad pathways. In this context, polyphenols have emerged as potential modifiers of human health, including immune health and longevity. However, beyond antioxidant effects, the specifics of *how* polyphenols and foods containing high levels of polyphenols and other phytochemicals impact health have remained relatively poorly interrogated.

Recent publications suggest the role of a polyphenol-rich diet in modulating epigenetics through direct gene methylation and through differential transcription of genes related to epigenetic regulation (Hoffman et al., 2023).

In this 90-day study, we provide some of the first clinical evidence suggesting that combinations of polyphenols and phytonutrients occurring naturally in Tartary buckwheat may have multiple effects on markers of longevity and immune system makeup.

By comparing participant’s epigenetic analyses before and after the 90-day intervention period, we were able to observe statistically significant changes in rate of aging as measured by the PCPhenoAge and PCGrimmAge and OmicAge algorithms in subgroups experiencing higher and lower rates of aging, respectively. This suggests the potential for the combination of nutrients administered to exert an effect on aging parameters in immune cells.

In analysis of immune cell subtypes using deconvolution methods, there were significant trends noted in subset analyses, but not across the aggregated data. As a representation of subset-specific data (reviewed above in **Table 5** above, in the PCPhenoAge evaluation), these included:

- Significant increases in CD4 T memory cells (PCPhenoAge – 1SD Higher) and CD8 T memory cells (PCPhenoAge – 1SD Higher)
- Significant decreases in B naive cells (PCPhenoAge – 1SD Lower)
- Significant decreases in Natural Killer cells (PCPhenoAge – Within 1 SD)

**Table 5:** Representation of changes in deconvoluted immune cell subsets using Epigenetic Age Accelerations (PCPhenoAge used as example).

|  |  |  |  |  |
| --- | --- | --- | --- | --- |
|  | Week 0 | Week 8 | Wilcoxon – P-Value | Clock Comparison |
| CD4Tmem | 0.096 | 0.095 | 0.934 | PCPhenoAge – Within |
| CD8Tmem | 0.053 | 0.052 | 0.427 | PCPhenoAge – Within |
| CD4Tnv | 0.073 | 0.077 | 0.117 | PCPhenoAge – Within |
| CD8Tnv | 0.034 | 0.032 | 0.178 | PCPhenoAge – Within |
| Bmem | 0.020 | 0.019 | 0.645 | PCPhenoAge – Within |
| Bnv | 0.042 | 0.040 | 0.470 | PCPhenoAge – Within |
| Treg | 0.008 | 0.009 | 0.427 | PCPhenoAge – Within |
| Baso | 0.019 | 0.018 | 0.594 | PCPhenoAge – Within |
| Eos | 0.012 | 0.009 | 0.786 | PCPhenoAge – Within |
| **NK** | **0.060** | **0.052** | **0.046** | **PCPhenoAge – Within** |
| Neu | 0.521 | 0.540 | 0.441 | PCPhenoAge – Within |
| Mono | 0.063 | 0.057 | 0.220 | PCPhenoAge – Within |
| CD4Tmem | 0.105 | 0.087 | 0.156 | PCPhenoAge – 1SD Lower |
| CD8Tmem | 0.059 | 0.045 | 0.219 | PCPhenoAge – 1SD Lower |
| CD4Tnv | 0.143 | 0.118 | 0.109 | PCPhenoAge – 1SD Lower |
| CD8Tnv | 0.055 | 0.039 | 0.078 | PCPhenoAge – 1SD Lower |
| Bmem | 0.019 | 0.017 | 0.297 | PCPhenoAge – 1SD Lower |
| **Bnv** | **0.067** | **0.055** | **0.031** | **PCPhenoAge – 1SD Lower** |
| Treg | 0.013 | 0.011 | 0.469 | PCPhenoAge – 1SD Lower |
| Baso | 0.018 | 0.014 | 0.016 | PCPhenoAge – 1SD Lower |
| Eos | 0.007 | 0.003 | 0.402 | PCPhenoAge – 1SD Lower |
| NK | 0.063 | 0.057 | 0.469 | PCPhenoAge – 1SD Lower |
| Neu | 0.398 | 0.512 | 0.109 | PCPhenoAge – 1SD Lower |
| Mono | 0.052 | 0.042 | 0.375 | PCPhenoAge – 1SD Lower |
| **CD4Tmem** | **0.071** | **0.107** | **0.031** | **PCPhenoAge – 1SD Higher** |
| **CD8Tmem** | **0.037** | **0.055** | **0.031** | **PCPhenoAge – 1SD Higher** |
| CD4Tnv | 0.044 | 0.063 | 0.063 | PCPhenoAge – 1SD Higher |
| CD8Tnv | 0.034 | 0.042 | 0.156 | PCPhenoAge – 1SD Higher |
| Bmem | 0.014 | 0.020 | 0.031 | PCPhenoAge – 1SD Higher |
| Bnv | 0.032 | 0.039 | 0.156 | PCPhenoAge – 1SD Higher |
| Treg | 0.007 | 0.005 | 0.438 | PCPhenoAge – 1SD Higher |
| Baso | 0.014 | 0.019 | 0.156 | PCPhenoAge – 1SD Higher |
| Eos | 0.002 | 0.007 | 0.201 | PCPhenoAge – 1SD Higher |
| NK | 0.029 | 0.043 | 0.063 | PCPhenoAge – 1SD Higher |
| Neu | 0.654 | 0.548 | 0.063 | PCPhenoAge – 1SD Higher |
| Mono | 0.061 | 0.051 | 0.563 | PCPhenoAge – 1SD Higher |

**TABLE 6:**
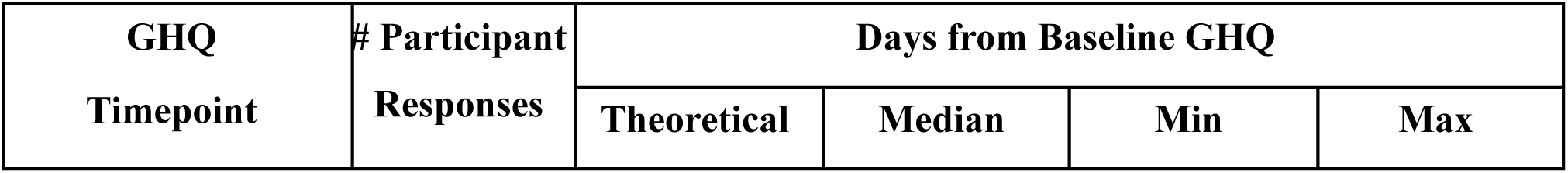
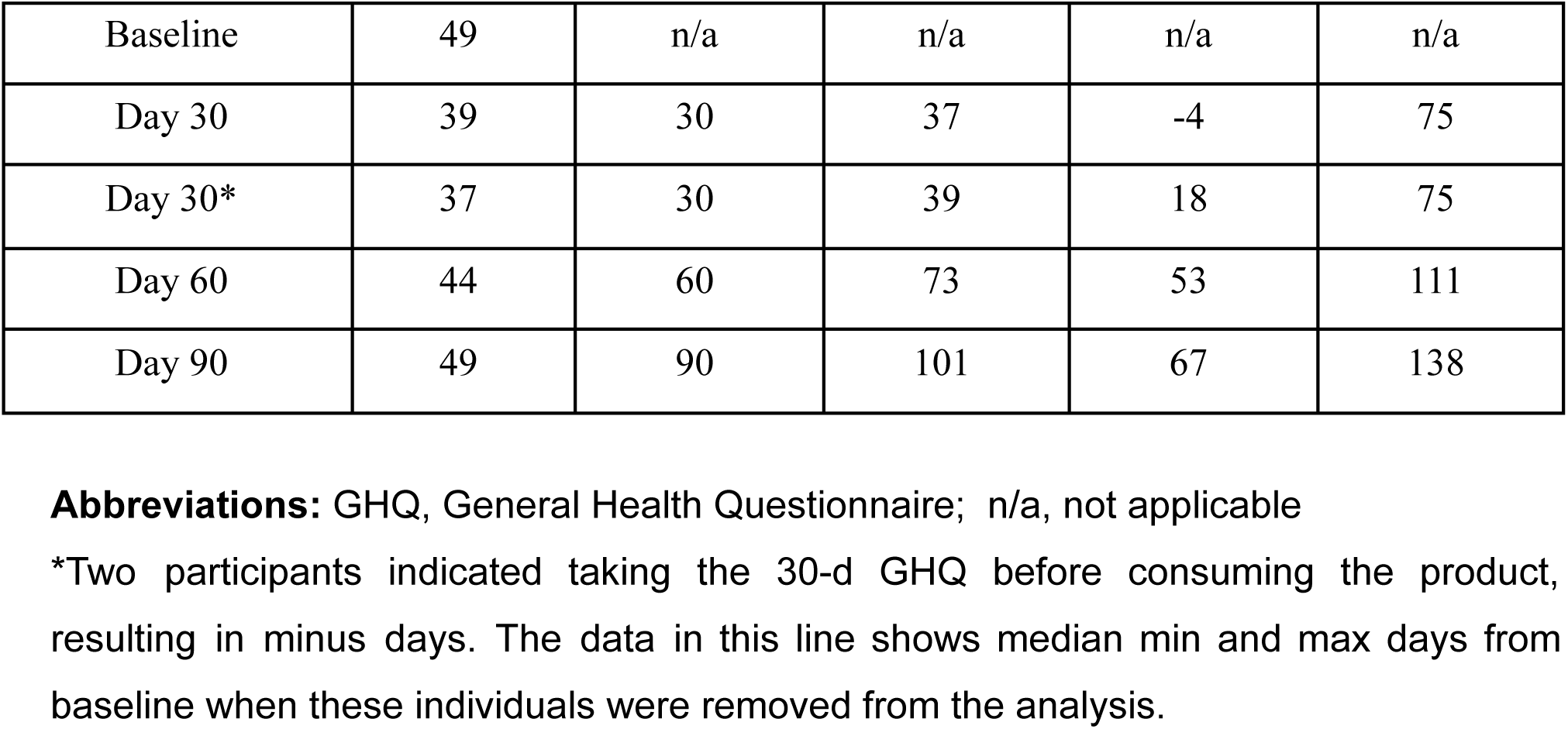
Time between GHQ Responses.

While the PCPhenoage 1SD higher population saw a decrease in speed of epigenetic age progression, this group also demonstrated an increase in CD4T and CD8T memory cells. Conversely, those starting the study at a lower overall epigenetic age score saw a decrease in naive B cells. These alterations in adaptive immune cells speak to potential effects on immune phenotypes over a 90 day interventional window.

Using genome-wide EWAS analysis comparing blood samples from the start and end of the study period, we identified 742 differentially methylated CPG sites at a p value of <0.001 with 242 hypermethylated and 480 hypomethylated sites controlled for potential overfitting. These differentially methylated sites were then analyzed using the GREAT software to identify GO pathways.

When we mapped CpG methylation against known biological processes using the GREAT software, a total of 22 pathways were linked to hypermethylated CpGs, while 134 were linked to hypomethylated CpGs. On review of these processes, the most highly enriched changes in biological pathways occurred within the hypermethylated CpG sites, where the largest fold enrichment (22x) was linked to ceramide kinase activity. We additionally found a 6.7-fold enhancement in COP9 activity. These relatively large effects, in contrast with the aforementioned immune cell deconvolution results, suggest that the more pronounced impact of the dietary supplementation occurred on upstream immune-related biological pathways measurable through epigenetics.

### Potential Implications of Enrichment of Ceramide Kinase Pathway Methylation

Ceramides are bioactive lipids found in plasma membranes regulating a host of cellular processes including cell cycle and immunity, in part through their effects as second messengers (Hilvo et al., 2020; Albeituni et al., 2019). Multiple studies show correlations between levels of circulating ceramides and risk for cardiometabolic and immunological dysfunction (Hilvo et al., 2020, de Mello et al., 2009).

Higher circulating ceramide levels are linked to cellular stressors including quality of diet. For example, consumption of palmitic acid elevated ceramide levels while the inverse occurred with polyunsaturated fat intake (Rosqvist et al., 2019).

Ceramide kinase (CERK) is an enzyme which adds phosphate to ceramide to generate ceramide-1-phosphate (C1P) (Al-Rashed et al., 2021). In humans, this enzyme is especially expressed in hepatic, renal, muscular and nervous tissue, and is present in plants and diverse animal species (Sugiura et al., 2002). C1P in turn helps regulate immune parameters, especially metabolism-associated inflammation, and has been strongly linked to pro-inflammatory signaling as well as the potential for anti-inflammatory signaling depending on cell type (Al-Rashed et al., 2021; Presa et al., 2016). C1P activity is believed to inhibit wound repair by promoting inflammation (Maus et al., 2022).

Activity of CERK in monocytes contributes to generation of inflammatory cytokines including IL-1 and monocyte chemoattractant protein-1 (MCP-1) as well as activation of the pro-inflammatory transcription factor NF-κB while inhibition of this pathway has the opposite effect (Al-Rashed et al., 2021). In a mouse model, CERK inhibition blocked inflammatory signaling in adipose tissue as well as suppressing diet-related weight gain and enhancing glucose metabolism (Mitsutake et al., 2012). CERK is also believed to promote mast cell degranulation (Vaquer et al., 2023).

CERK is also a known regulator of cell cycle arrest (senescence), an established contributor to age-related diseases as well as inflammation. Inhibition of CERK has been demonstrated in preclinical data to reduce senescent cell burden, while C1P activity has been shown to block apoptosis and enhance cell survival (Millner et al., 2022; Gómez-Muñoz, 2006).

In preclinical work, application of polyphenolics has been shown to modulate ceramide pathway outcomes. For example, rutin administration has been found to suppress renal ceramide levels in a mouse study (Ma et al., 2018). Additionally, observational data in humans find lower levels of circulating ceramides in those with adherence to the Mediterranean diet (Yang and Chen, 2022).

In a network pharmacology analysis of the compounds present in Tartary buckwheat, RAF1 was identified as a key target. RAF1 (RAF proto-oncogene serine/threonine-protein kinase) has a known role in cellular proliferation (Lu et at., 2017). RAF1 in turn is known to be phosphorylated by the downstream effects of ceramide (Yao, 1995).

The sum total of this data suggest the potential for the effects of Tartary buckwheat on immune, longevity and epigenetic outcomes to aggregate around an anti-senescence signal by way of ceramide kinase pathway inhibition.

### Potential Implications of Enrichment of COP9 Signalosome Pathway Methylation

Regarding COP9, this is a conserved protein complex composed of 9 subunits found across eukaryotic cells, and well described in plants and animals. In plants, the COP9 signalosome regulates gene expression and resilience to abiotic stress. COP9 is an established cell-cycle regulator through impacts on ubiquitination and transcriptional modification (Doronkin et al., 2003). Genes associated with the COP9 signalosome are linked to regulation of senescence (Leal et al., 2008). The COP9 signalosome has more recently been implicated in the modulation of neuroinflammation including effects on microglial cells, and appears to play a key role in maturation of the autophagosome (Tian et al., 2023; Su et al., 2011) It has previously been shown that curcumin polyphenols are capable of impacting COP9 (Lim et al., 2016).

### Changes Across Diverse Immune and Longevity Pathways Suggest Specific Role for Tartary Buckwheat in “Food is Medicine” Discussion

Suboptimal diet has been clearly established as a major risk factor for global death and disability (Afshin et al., 2019). However, recommendations around dietary modification are largely generalized (e.g., eat more fruits, vegetables and whole grains). While this may help at a population level in preventing disease, there has also been a call to understand whether certain foods and nutrients (beyond total calories, macro and micronutrients) may serve to provide an outsized benefit in the health of the individual. As immune-mediated or related conditions are among the most common chronic diseases, dietary interventions targeting immune dysfunction represent an important part of the conversation of “food is medicine.”

The data from this pilot study helps provide a more nuanced understanding of how combinations of naturally-occurring phytochemicals might induce beneficial effects on humans through activation of biological pathways involved in immune modulation and immune aging. Compared to pharmaceuticals operating with high potency effects on receptors, the biological impact of naturally-occurring nutrients like polyphenols at levels similar to what has typically been found in diets of long-lived populations around the world is expected to be more gradual in onset and less pronounced in effect. Here, we provide preliminary data demonstrating the impact of select phytochemicals from Tartary Buckwheat in effecting changes across biological pathways involved in health and disease.

## Conclusion

In this study, we present novel data indicating the potential for phytochemical nutrients found within Tartary buckwheat to act on multiple metrics of epigenetic immune age, immune cells and related cellular pathways. The polyphenol-rich supplement designed around key bioactive nutrients naturally occurring in Tartary buckwheat appeared to directionally influence CpG methylation patterns in subsets of participants with higher and lower rates of biological aging as measured by the PCPhenoAge, PCGrimAge and OmicAge aging algorithms. These changes were correlated with changes in peripheral immune cell patterns. Changes in GO pathways additionally suggest the potential for effects on multiple immune and cellular regulatory mechanisms, especially those related to ceramide kinase. These results suggest that polyphenols and associated bioactive nutrients naturally occurring in Tartary buckwheat may significantly influence epigenetic age measurements that may be driven by or reflected in changes in the immune system and associated pathways.

## Author Contributions

Conceptualization of the experiment was performed by JSB, AP and RS. Clinical visits, compliance checks and recruitment of participants performed by AC and SSM. Processing of samples performed by TLM. Statistical analysis was performed by VBD. Interpretation of data and writing of manuscript performed by AP, JSB and VBD.

## Data Availability

All data produced in the present study are available upon reasonable request to the authors

## Acknowledgements

Special thanks to DeAnn Liska for guidance in developing the study design and assistance in reporting the clinical data. Additionally, we appreciate the contributions of Sierra Stromberg in participant follow up and data management, as well as Annie Prestrud in coordinating data collection.

## Data Sharing Statement

The data that support the findings of this study are available from the corresponding authors in the context of reasonable request.

## Trial Registration

ClinicalTrials.gov Identifier: NCT05234203

## Supplementary Files

**Supplementary File 1:** Contains Appendix 1 (Population Selection Summary Chart), Appendix 2 (GHQ Questionnaire), and Appendix 3 (GHQ Individual Question Results for PP Population).

**Supplementary File 2:** Contain a table noting the full list of differentially methylated loci (DML) identified in the comparison between the end of the trial (day 90) and baseline (day 0). Information reported in the table is as follows: Column A) CpG probeset identified as significantly methylated. Column B) fold change (log normalized), where values greater 0 are representative of a methylation change higher at the final timepoint compared to baseline, and vice versa. Column C) the moderated t-statistic calculated from the statistical test implemented; Column D) the unadjusted p-value; Column E) the adjusted p-value based on a False Discovery Rate adjustment; and F) the Gene ID associated with the probe ID.

**Supplementary File 3:** The full list of gene ontology terms identified for the DMLs outputted from the GREAT software. Gene ontology (GO) terms identified with the hypermethylated DMLs, CpGs identified as logFC > 0 in Supplementary File 2, are noted in the first sheet labeled as “Hypermethylated”, while hypomethylated DMLs, CpGs identified as logFC < 0 in Supplementary File 2, are noted in the second sheet labeled as “Hypomethylated”. Important columns include: Column A) discriminates whether the gene ontology reported is a Biological Process (BP), Molecular Function (MF), or a Cellular Component (CC) term; Column B) the specific GO term ID; Column G) the fold enrichment, and Column I) the raw p-value.

