## Supplementary File 1 for "The Impact of a Polyphenol-Rich Supplement on Epigenetic and Cellular Markers of Immune Age: A Pilot Clinical Study"

### Contents: Appendix 1-3

#### **Appendix 1:** Population Selection Summary Chart

| Study ID | COVID Status | Include in ITT | Include in PP | Rationale |
|---|---|---|---|---|
| BBH-008-0 004 | No | YES | YES | Added a supplement that could impact questionnaire data |
| BBH-009-0 012 | No | YES | YES | Added a supplement that could impact questionnaire data |
| BBH-010-0 001 | No | YES | YES | |
| BBH-012-0 009 | No | YES | YES | |
| BBH-013-0 013 | No | YES | YES | |
| BBH-014-0 007 | No | YES | YES | |
| BBH-015-0 010 | YES | YES | YES | |
| BBH-016-0 002 | YES | YES | YES | |
| BBH-017-0 003 | No | YES | NO | Added a curcumin supplement |
| BBH-018-0 005 | No | YES | YES | |
| BBH-019-0 008 | No | YES | NO | Low HTB compliance |
| BBH-020-0 011 | No | YES | YES | |
| BBH-022-0 014 | No | YES | YES | |
| BBH-023-0 025 | No | YES | YES | |
| BBH-024-0 016 | No | NO | NO | Did not complete, no 2$^{nd}$ lab test |
| BBH-027-0 018 | No | YES | YES | |
| BBH-028-0 017 | No | YES | YES | |

| | | | | |
|---|---|---|---|---|
| BBH-029-0021 | No | YES | YES | |
| BBH-030-0023 | No | YES | YES | |
| BBH-031-0020 | YES | YES | YES | |
| BBH-033-0019 | YES | YES | YES | |
| BBH-036-0026 | YES | YES | YES | |
| BBH-037-0027 | No | YES | YES | |
| BBH-038-0028 | YES | YES | YES | |
| BBH-039-0030 | No | YES | NO | Low HTB compliance |
| BBH-040-0029 | No | YES | YES | |
| BBH-041-0031 | YES | YES | YES | |
| BBH-042-0033 | No | YES | YES | |
| BBH-043-0032 | No | NO | NO | Did not complete, no 2nd lab test |
| BBH-044-0034 | No | YES | NO | Low HTB compliance |
| BBH-046-0036 | No | YES | YES | |
| BBH-047-0037 | YES | YES | YES | |
| BBH-048-0038 | YES | YES | YES | |
| BBH-049-0040 | YES | YES | YES | |
| BBH-050-0039 | YES | YES | YES | |
| BBH-051-0043 | No | YES | YES | |
| BBH-052-0041 | YES | YES | YES | |
| BBH-053-0042 | YES | YES | NO | Low HTB Compliance |
| BBH-054-0044 | No | YES | YES | |
| BBH-055-0045 | No | YES | YES | |

| | | | | |
|---|---|---|---|---|
| BBH-057-0 047 | No | YES | NO | Post-menopausal and started estriol |
| BBH-058-0 048 | No | YES | NO | Added supplements and low HTB compliance |
| BBH-059-0 050 | No | YES | YES | |
| BBH-060-0 049 | No | YES | YES | |
| BBH-065-0 052 | No | YES | YES | |
| BBH-066-0 053 | No | YES | YES | |
| BBH-067-0 054 | YES | YES | YES | |
| BBH-068-0 056 | No | YES | YES | |
| BBH-069-0 057 | No | YES | YES | |

### **Appendix 2:** GHQ Questionnaire

Instructions: These questions are about how you feel and how things have been with you **during the past 4 weeks.** For each question, please give the one answer that come closest to the way you have been feeling. Choose only one option for each question.

1.Over the past 4 weeks, would you say your health is:

- ○ Excellent
- ○ Very Good
- ○ Good
- ○ Fair
- ○ Poor

2. Considering how you were feeling prior to the past month, would you say that your overall health has changed?

- ○ Yes – Much better over the last 4 weeks
- ○ Yes – Somewhat better over the last 4 weeks
- ○ No – About the same as before
- ○ Yes – Somewhat worse over the last 4 weeks
- ○ Yes – Much worse over the last 4 weeks

3. Considering gastrointestinal effects, such as reflux, heartburn, stomach rumbling, bloating, intestinal pain and discomfort, and flatulence, please comment on your experience over the past 4 weeks.
    - ○ Much worse (higher number) and/or more intense symptoms than usual
    - ○ Somewhat worse than usual (number and/or intensity)
    - ○ No change in symptoms compared to usual
    - ○ Somewhat fewer and/or less intense symptoms than usual
    - ○ Much less in number and/or less intensity than usual

4. Have you had changes to your bowel habits (frequency of going to the bathroom, comfort of bowel movement, form of stool (e.g., loose and watery or very hard) over the past 4 weeks?
    - ○ Much worse than usual
    - ○ Somewhat worse than usual
    - ○ About the same as usual
    - ○ Somewhat better than usual
    - ○ Much better than usual

5. Over the past 4 weeks, have you experienced brain fog?
    - ○ Not at all
    - ○ Infrequently
    - ○ Occasionally
    - ○ Often
    - ○ Very often

6. Considering the health of your skin over the last 4 weeks, have you experienced rashes, felt your skin is itchy or otherwise unhealthy?
    - ○ Not at all
    - ○ Infrequently
    - ○ Occasionally
    - ○ Often
    - ○ Very often

7. If you have skin symptoms, were these:
    - ○ Much worse than usual

- ○ Somewhat worse than usual
- ○ About the same as usual
- ○ Somewhat better than usual
- ○ Much better than usual

8. Over the past 4 weeks, other than an actual cold or illness, have you experienced Upper Respiratory Tract (URT) symptoms, such as runny nose, congestion or sneezing?
   - ○ Not at all
   - ○ Infrequently
   - ○ Occasionally
   - ○ Often
   - ○ Very often

9. Over the past 4 weeks, would you say your Upper Respiratory Tract (URT) symptoms were?
   - ○ Much worse than usual
   - ○ Somewhat worse than usual
   - ○ About the same as usual
   - ○ Somewhat better than usual
   - ○ Much better than usual

10. When considering your energy levels over the past 4 weeks, would you say you had a lot of energy?
    - ○ All of the time
    - ○ Most of the time
    - ○ Some of the time
    - ○ A little of the time
    - ○ None of the time

11. Have you felt like your energy levels have changed over the last 4 weeks?
    - ○ Much less energy than usual
    - ○ Somewhat less energy than usual
    - ○ About the same as usual
    - ○ Somewhat more energy than usual
    - ○ Much more energy than usual

12. Over the past 4 weeks, have you experienced low mood?
    - Very often
    - Often
    - Occasionally
    - Infrequently
    - Not at all

13. Over the past 4 weeks, have you felt in control of your health?
    - Not at all
    - Infrequently
    - Occasionally
    - Often
    - Very often

14. During the past 4 weeks, how often have you had trouble sleeping?
    - Not at all
    - Infrequently
    - Occasionally
    - Often
    - Very often

15. Please tell us about how your sleeping experience has been over the past 4 weeks, compared to your usual sleeping experience.
    - Much less sleep than usual
    - Somewhat less sleep than usual
    - About the same sleep as usual
    - Somewhat more sleep than usual
    - Much more sleep than usual

### **Appendix 3:** GHQ Individual Question Results for PP Population*

| GHQ Question<br>Over prior 4 wks | Scoring | Statistic | Visit 1<br>Day 0 | Visit 4<br>Day 30 | Visit 5<br>Day 60 | Visit 6<br>Day 90 |
|---|---|---|---|---|---|---|

| | | # Completed | *36* | *29†* | *33†* | *36* |
|---|---|---|---|---|---|---|
| ***Question 1:*** General Health rating | Poor=1, Fair=2, Good=3, Very Good=4, Excellent=5 | Mean (SD) | 4.0 (0.8) | 4.0 (0.8) | 4.0 (0.7) | 3.7 (1.0) |
| | | Median (min, max) | 4.0 (2.0, 5.0) | 4.0 (2.0, 5.0) | 4.0 (2.0, 5.0) | 4.0 (2.0, 5.0) |
| **Question 2:** Change in General Health | Much worse=1, Some worse=2, Same=3, Some better=4, Much better=5 | Mean (SD) | 3.2 (0.5) | 3.1 (0.4) | 3.3 (0.6) | 2.8 (0.6) |
| | | Median (min, max) | 3.2 (2.2, 4.0) | 3.0 (2.0, 4.0) | 3.0 (2.0, 5.0) | 3.0 (1.0, 4.0) |
| **Question 3:** Status of GI symptoms | Much worse=1, Some worse=2, Same=3, Some better=4, Much better=5 | Mean (SD) | 2.9 (0.4) | 2.9 (0.9) | 2.9 (0.7) | 2.9 (0.6) |
| | | Median (min, max) | 3.0 (1.0, 4.0) | 3.0 (1.0, 5.0) | 3.0 (1.0, 4.0) | 3.0 (1.0, 4.0) |
| **Question 4:** Change in Bowel Habits | Much worse=1, Some worse=2, Same=3, Some better=4, Much better=5 | Mean (SD) | 2.9 (0.3) | 2.8 (0.6) | 2.9 (0.5) | 2.8 (0.5) |
| | | Median (min, max) | 3.0 (2.0, 3.0) | 3.0 (2.0, 5.0) | 3.0 (2.0, 4.0) | 3.0 (1.0, 4.0) |
| **Question 5:** Experience of Brain Fog | Very Often=1, Often=2, Occasion=3, Infrequent=4, None=5 | Mean (SD) | 4 (0.8) | 4.3 (0.7) | 4.1 (0.8) | 4.1 (0.8) |
| | | Median (min, max) | 4.0 (2.0, 5.0) | 4.0 (3.0, 5.0) | 4.0 (2.0, 5.0) | 4.0 (3.0, 5.0) |
| **Question 6:** Experience of Unhealthy Skin symptoms (rash/itch) | Very Often=1, Often=2, Occasion=3, Infrequent =4, None=5 | Mean (SD) | 4.5 (0.9) | 4.8 (0.5) | 4.5 (0.9) | 4.6 (0.7) |
| | | Median (min, max) | 5.0 (2.0, 5.0) | 5.0 (3.0, 5.0) | 5.0 (1.0, 5.0) | 5.0 (3.0, 5.0) |
| **Question 7:** Skin Symptom Change Rating | Much worse=1, Some worse=2, Same=3, Some better=4, Much better=5 | Mean (SD) | 2.9 (0.5) | 3.0 (0.4) | 3.0 (0.4) | 2.9 (0.4) |
| | | Median (min, max) | 3.0 (1.0, 4.0) | 3.0 (2.0, 4.0) | 3.0 (2.0, 4.0) | 3.0 (2.0, 4.0) |
| **Question 8**: Presence of Non-URT | Very Often=1, Often=2, Occasion=3, | Mean (SD) | 4.4 (1.1) | 4.3 (0.9) | 4.2 (1.1) | 4.3 (1.1) |
| | | Median (min, max) | 5.0 (1.0, 5.0) | 5.0 (2.0, 5.0) | 5.0 (2.0, 5.0) | 5.0 (1.0, 5.0) |

| | | | | | | |
|---|---|---|---|---|---|---|
| (cold/illness) Symptoms | Infrequent=4, None=5 | | | | | |
| **Question 9:** Change in URT Symptoms | Much worse=1, Some worse=2, Same=3, Some better=4, Much better=5 | Mean (SD) | 2.9 (0.5) | 2.9 (0.5) | 2.8 (0.6) | 2.9 (0.5) |
| | | Median (min, max) | 3.0 (1.0, 4.0) | 3.0 (2.0, 4.0) | 3.0 (1.0, 4.0) | 3.0 (1.0, 4.0) |
| **Question 10:** Experienced "a lot" of Energy | None=1, A Little=2, Some=3, Most=4, All=5 | Mean (SD) | 3.3 (0.8) | 3.4 (0.8) | 3.4 (0.8) | 3.3 (0.8) |
| | | Median (min, max) | 3.5 (1.0, 4.0) | 4.0 (1.0, 4.0) | 4.0 (1.0, 4.0) | 3.0 (1.0, 5.0) |
| **Question 11:** Change in Energy Level | Much Less=1, Some Less=2, Same=3, Some More=4, Much More=5 | Mean (SD) | 2.8 (0.6) | 3.1 (0.5) | 3.1 (0.6) | 2.8 (0.7) |
| | | Median (min, max) | 3.0 (1.0, 4.0) | 3.0 (2.0, 4.0) | 3.0 (2.0, 4.0) | 3.0 (1.0. 4.0) |
| **Question 12:** Experienced Low Mood | Very Often=1, Often=2, Occasion=3, Infrequent=4, None=5 | Mean (SD) | 3.8 (1.0) | 4.0 (1.0) | 3.8 (1.0) | 3.9 (0.8) |
| | | Median (min, max) | 4.0 (2.0, 5.0) | 4.0 (2.0, 5.0) | 4.0 (1.0, 5.0) | 4.0 (2.0, 5.0) |
| **Question 13:** Feelings of being in Control of Health | None=1, Infrequent=2, Occasion=3, Often=4, Very Often=5 | Mean (SD) | 4.2 (0.9) | 4.2 (0.8) | 3.8 (1.0) | 4.0 (0.9) |
| | | Median (min, max) | 4.0 (1.0, 5.0) | 4.0 (3.0, 5.0) | 4.0 (1.0, 5.0) | 4.0 (2.0, 5.0) |
| **Question 14:** Experience of Trouble Sleeping | Very Often=1, Often=2, Occasion=3, Infrequent=4, None=5 | Mean (SD) | 3.6 (1.0) | 3.7 (0.8) | 3.9 (0.9) | 3.7 (0.7) |
| | | Median (min, max) | 3.5 (1.0, 5.0) | 4.0 (2.0, 5.0) | 4.0 (2.0, 5.0) | 4.0 (2.0, 5.0) |
| **Question 15:** Change in Sleep from Usual Sleeping Pattern | Much Less=1, Some Less=2, Same=3, Some More=4, Much More=5 | Mean (SD) | 2.7 (0.5) | 3.1 (0.5) | 2.9 (0.4) | 2.7 (0.5) |
| | | Median (min, max) | 3.0 (2.0, 4.0) | 3.0 (2.0, 5.0) | 3.0 (2.0, 4.0) | 3.0 (1.0, 3.0) |

**Abbreviations:** d, day; GI, gastrointestinal; max, maximum; min, minimum; SD, standard deviation; URT, upper respiratory tract; wks, weeks

*The data also excludes two individuals consuming supplements that were likely to affect the results as well as two individuals who had baseline timing that were not feasible
